# Short-term changes in objectively measured activity predict brain atrophy and disability progression in multiple sclerosis

**DOI:** 10.1101/2025.01.27.25321205

**Authors:** Kathryn C. Fitzgerald, Muraleetharan Sanjayan, Blake Dewey, Pratim Guha Niyogi, Nicole Bou Rjeily, Yasser Fadlallah, Alice Delaney, Alexandra Zambriczki Lee, Safiya Duncan, Chelsea Wyche, Ela Moni, Peter A. Calabresi, Vadim Zipunnikov, Ellen M. Mowry

**Affiliations:** Division of Neuroimmunology and Neurological Infections, Department of Neurology, Johns Hopkins School of Medicine, Baltimore, MD; Department of Biostatistics, Johns Hopkins Bloomberg School of Public Health, Baltimore, MD

**Keywords:** multiple sclerosis, neuroimaging, wearables, accelerometry

## Abstract

**Objective:** To evaluate how within-person changes in accelerometry-derived activity patterns translate to brain atrophy and disability worsening in people with multiple sclerosis (PwMS).

**Methods:** We included PwMS aged ≥40 years with approximately annual brain MRI who wore GT9X Actigraph accelerometers every three months over three years. Accelerometry-derived indices included total and 2-hour specific activity, sedentary time, and circadian rhythm parameters. Confirmed disability worsening was characterized using the composite Expanded Disability Status Scale-plus (EDSS+) and whole brain segmentation used SLANT-CRUISE. We modeled within- and between-person effects using Cox (for EDSS+) and linear mixed effects (for MRI) multivariable-adjusted models adjusted for age, sex, and body mass index that included only accelerometry measures obtained prior to EDSS+.

**Results:** Among 239 PwMS (mean age 54.8, 29% male), 120 experienced EDSS+-confirmed progression over a mean 2.9 years (SD: 1.1 years). Participants wore accelerometers an average of 7.4 times over 67 days. Total activity declined an average of 48,694 activity counts (~0.10 SD per year; 95% CI: −33092, −64297; p=1.21×10^−9^), and people with progressive MS exhibited more pronounced declines in total activity relative to RRMS (p=2.34×10^−5^). Within-person decreases in daytime activity (particularly 8:00-14:00) were significantly associated with higher risk of EDSS+. For example, a 1 SD decrease in within-person (individual-level) activity from 8:00-10:00, 10:00-12:00 and 12:00-14:00 was associated with a respective 1.20 (95% CI: 1.04, 1.39; p=0.01), 1.22 (95% CI: 1.04, 1.41; p=0.008), and 1.23 (95% CI: 1.07, 1.42; p=0.007) higher risk of confirmed disability progression by EDSS+. MRI also models demonstrated that within-person declines in morning activity (8:00-10:00) were associated with greater whole brain, deep gray matter and thalamic volume loss (for whole brain −0.16%; 95% CI: −0.29, −0.04; p=0.009; deep gray: −0.32%; 95% CI: −0.13, −0.51; p=0.0009; thalamic: −0.30%; 95% CI: −0.52, −0.08; p=0.007). Lower between-person mean MVPA was associated with lower brain volumes over time but was not associated with EDSS+.

**Interpretation:** Within-person reductions in daytime activity patterns precede clinical disability worsening and brain atrophy in PwMS. Longitudinal accelerometry may offer sensitive, non-invasive biomarkers of subclinical disease progression in MS.

## Introduction

Multiple sclerosis (MS) is the most common cause of non-traumatic disability in young adults and affects millions of people worldwide.^1,2^ Most people with MS are initially diagnosed with relapsing-remitting MS (RRMS), in which immune-mediated demyelinating attacks are common. Approximately half of people with RRMS will transition to a progressive phase of the disease, typically around age 45, where disability accumulation becomes more apparent with a gradual worsening of physical and cognitive functioning.^3^ A small percentage of PwMS (~10%) have progressive-onset disease, in which steady deterioration of function is apparent from diagnosis.^2,3^ Notably, while current disease-modifying MS therapies are effective in reducing immune-mediated attacks, they do not meaningfully slow insidious progression of MS, thought to be occurring as a result of underlying neurodegeneration.^4,5^ These processes likely begin years prior to the manifestation of clinical symptoms. Thus, there remains a critical need to develop therapies that can effectively target this phase of the disease.

One current roadblock in the path to advancing progressive MS therapies remains a lack of accurate measures of clinical disease progression.^6^ The Expanded Disability Status Scale (EDSS), widely used in clinical trials, has significant limitations due to its semi-quantitative and non-linear nature, which affects its reliability and responsiveness.^7,8^ These drawbacks translate to larger sample sizes and longer follow-ups in clinical trials, increasing costs and complicating drug development. Additionally, the EDSS may fail to identify individuals in early progressive stages, particularly those with suspected clinical worsening that cannot be confirmed over short intervals, leaving them classified as having RRMS and ineligible to participate in progressive MS trials. Improved methods to track progression are essential to overcome these challenges.

Wearable technologies, such as accelerometry, provide an opportunity to better detect and monitor disability progression.^9–12^ Accelerometry offers objective, real-world data on physical activity, circadian rhythms, and sleep patterns and is relatively cheap and accessible.^13,14^ Short-term, continuous monitoring allows for insight into variation in activity within a 24-hour period or over the course of several days, indicative of disease progression that may be missed during a traditional clinical visit.^10^ Furthermore, as the use of telemedicine continues to grow, real-world activity data from accelerometers could help supplement virtual care; patients with concerning activity patterns could be prioritized for in-person visits, while stable individuals could continue with hybrid care models.^15,16^ By integrating accelerometry into routine care and clinical trials, MS monitoring and management could become more accurate, efficient, and likely equitable.

However, to date, most wearable studies in MS have been (1) cross-sectional, (2) primarily focused on the correlation of total step count with symptoms or disability outcomes like the EDSS, and (3) have considered group (or average)-level effects.^9,11,12,17,18^ To our knowledge, no studies have included potentially more sensitive outcomes like those derived from brain MRI (e.g., total brain or gray matter volume) or have incorporated changes in activity at the individual level as predictors of brain atrophy or changes in clinical outcomes in MS. For example, findings linking sensitive measures of activity (or changes therein) over the course of the day to gray matter loss are particularly significant, as such atrophy is strongly predictive of worse long-term disability in PwMS.^19^ Furthermore, consideration of individual-level effects is crucial for advancing precision medicine approaches in MS, as it can inform early intervention or tailored disease monitoring strategies. Here, we evaluated how measures derived from accelerometry, including at specific times of the day and overall activity pattern, change over time and how individual-level changes in these measures translate to brain compartment atrophy and disability progression.

## Materials and Methods

### Study population

We included participants from the Home-based Evaluation of Actigraphy to predict Longitudinal Function in MS (HEAL-MS) cohort, as described previously. Briefly, HEAL-MS participants were recruited from the Johns Hopkins MS Precision Medicine Center of Excellence between January 2021 and March 2023 and were aged 40 years or older, did not have known significant comorbidities that could impair physical activity (e.g., heart failure, end-stage renal disease), did not have a MS relapse within the six months prior to enrollment, and had a baseline EDSS score of 6.5 or lower. We developed these criteria to (1) ensure that among the relapsing-remitting MS (RRMS) subgroup, participants were age-wise are at an elevated risk for transitioning to the progressive phase of MS; (2) to ensure that changes in accelerometry or disability were likely to be related to changes in MS rather than other conditions; and (3) to avoid confounding for baseline scores that were impacted by recovery from a relatively recent relapse. We categorized individuals into three groups. One group included people with stable RRMS who had no evidence of suspected or confirmed progression. Another group included people with confirmed disability worsening on EDSS (i.e., progressive MS [PMS]). The final group included people with RRMS who were suspected of clinical progression but did not have sustained disability worsening. Here, confirmed disability worsening was defined as an EDSS increase of at least 1.0 points for individuals with baseline EDSS ≤ 5.5 or an increase of at least 0.5 points for individuals with baseline EDSS at least 6 that was sustained at least 24 weeks later. A neurologist specializing in MS reviewed electronic healthcare records (EHR) to confirm eligibility for each participant and to assign PwMS to individual groups. The two RRMS groups were matched for age (± 2 years), sex at birth, race/ethnicity, and efficacy class of current DMT. All participants provided written consent in accordance with the Declaration of Helsinki, and the study was approved by the Johns Hopkins Institutional Review Board.

### Accelerometry measures

HEAL-MS participants wore GT9X Actigraph accelerometers that feature a built-in tri-axial accelerometer at pre-specified 3-month intervals throughout the course of the study. At each wear, individuals were instructed to wear the device on the wrist of their non-dominant hand continuously for 24 hours per day over an approximate two-week period. Each device was configured such that it recorded three-dimensional acceleration at 30 Hz with a range of ±8 G. Raw acceleration data in .gt3x format were downloaded using ActiLife v6.134 Lite Edition,^20^ and binary activity data were processed using the read.gt3x package in R, converted into activity count data with 60-second epochs, and formatted for analysis with 1440 minutes per day (midnight to 11:59 PM).^12^ For each wear-period, we defined non-wear time as an interval of at least 90 minutes in which all minute-level activity counts were equal to 0. As in previous analyses, a valid day during a given wear-period was defined as those in which wear-time exceeded 90% of the day (i.e., at least 1296/1440 minutes).^21^ We excluded wear-periods in which an individual had at less than 3 valid wear-days (<1% of wear-times were removed).

For our primary analyses, we performed analyses examining log-transformed 2-hour specific activity counts. As in our prior analysis,^22^ we expect that the 2-hour specific windows would provide some granularity as to how a person’s activity changes over the course of the day, while also allowing us to estimate these changes reliably from an analytical perspective. For secondary analyses, we also calculated total activity levels in a 24-hour period as well as a series of measures summarizing different physical and circadian activity patterns. For physical activity, these included: (1) light intensity physical activity (LIPA); (2) moderate-to-vigorous physical activity (MVPA); (3) number of sedentary minutes; (4) measures of fragmentation including the sedentary to active transition probability (SATP) and (5) active to sedentary transition probability (ASTP).^23^ For circadian measures, we included parametric and non-parametric indices. Parametric indices were using extended cosinor models and include: (1) amplitude-half the distance between the peak/trough of activity; (2) acrophase-maximal activity point in cycle (3) mesor-mean value around which the cycle oscillates.^24^ Non-parametric indices included diurnal landmarks such as (1) M10-most active 10-hours of a 24-hour period; (2) L5-least active 5-hours of a 24-hour period; (3) daytime activity ratio (DARE), defined as activity from 8:00-20:00 divided by the total activity from 0:00-23:59; (4) relative amplitude defined as (M10-L5)/(M10+L5); (5) inter-daily stability (IS) in a 24-hour period; and inter-daily variability (IV) in a 24-hour period.^25,26^ For each wear-time, we averaged values across all days for each candidate measures.

### Outcomes

#### Clinical outcomes

In HEAL-MS, neurologic exams are performed every 6 months. EDSS exam was conducted after the fitting of the accelerometer and performed by a masked (blinded) EDSS-trained physician. A modified MS Functional Composite (MSFC), which includes the 9-hole peg test, the timed 25-foot walk test, and the Symbol Digit Modalities Test (SDMT), was performed by a masked, trained study team member. The primary clinical disability outcome for our study was the EDSS+. Progression by EDSS+ occurs due to EDSS change (change at any time point of >1.0 point if baseline EDSS is <5.5 or of >0.5 if baseline EDSS is ≥6.0, that is confirmed at a subsequent visit at least 6 months later) OR 20% worsening on either the timed 25-foot walk test (T25FW)^27^ or the nine-hole peg test (9HPT)^28^ that is confirmed at a subsequent visit ≥6 months later. Here, we considered ‘baseline’ using a roving EDSS reference value, as this approach has been shown to more efficiently identify progression or worsening disability events.^29^

#### MRI

Research MRIs obtained on a 3T Siemens scanner were acquired as a part of the HEAL-MS protocol at baseline and at the conclusion of Year 2. Interim MRIs collected under an identical protocol obtained as a part of clinical care were also included, when available. Our MRI processing pipeline was developed for a large-scale, multi-site MS clinical trial in which clinically acquired scans are analyzed that allowed us to increase power and better characterize longitudinal trajectories by including interim MRI scans obtained on multiple scanners. It was designed to handle various image sources, including T1-weighted, T2-weighted, PD-weighted, and T2-FLAIR imaging for processing, and images before and after contrast are usable (and a subset of contrasts can be missing).^30,31^ Multi-slice image volumes undergo deep learning-based super-resolution with SMORE (Synthetic Multi-Orientation Resolution Enhancement),^32–34^ followed by alignment of the super-resolved and 3D-acquired images using a longitudinal registration system. This system, using the advanced normalization tools (ANTs) software package, is optimized to manage diverse longitudinal data. The alignment process involves three main steps: (1) aligning each subject to a reference atlas; (2) adjusting images across different time points; and (3) aligning images within each time point. Rigid transformations are employed to maintain anatomical integrity. Applying Harmonization with Attention-based Contrast, Anatomy, and Artifact Awareness (HACA3), three contrast targets at each time point are generated: 3D T1-weighted MPRAGE, 3D T2-FLAIR, and multi-slice T2-weighted images, all of which attempt to adhere to standardized protocol from the Johns Hopkins MS on a single Siemens 3T scanner.^35,36^ The HACA3-harmonized images serve as inputs for white matter lesion segmentation using a 3D U-Net architecture. T1-weighted images, with any lesions filled, are segmented using Spatially Localized Atlas Network Tiles (SLANT), which segments the brain into 133 distinct regions of interest.^37,38^ After segmentation, the surface is reconstructed using Cortical Reconstruction Using Implicit Surface Evolution (CRUISE) to refine SLANT segmentation boundaries and measure cortical thickness.^39,40^ A U-Net-based algorithm is used to calculate the total intracranial volume as a surrogate for head size, and all MRI volumes are normalized for head size.

### Statistical analysis

We calculated descriptive statistics for accelerometry measures including means or medians along with standard deviations or interquartile ranges across demographic or clinical characteristics. Initial analyses also evaluated whether the rate of disability progression (using EDSS+) varied across HEAL-MS subgroups or by clinical phenotypes (e.g., progressive vs. relapsing) using a log-rank test and graphically using the complement of Kaplan Meier curves for cumulative incidence. We structured our analysis of accelerometry measures into two complementary components: (1) changes in accelerometry measures over follow-up and (2) how these changes translate to changes in outcomes. We considered each accelerometry measure (e.g., TAC, TAC_0:00-2:00,_ ASTP, among others) in separate models. For the first component, we assessed whether changes in candidate accelerometry biomarkers were detectable prior to confirmed disability progression and whether changes over time varied across different HEAL-MS or clinical subgroups using linear mixed effects (LME) models adjusted for age, sex, race and BMI (as kg/m^2^). Tests for differences in accelerometry measures over time across HEAL-MS subgroups were performed as the cross-product between time and subgroup. Then, for the second component of our analytic pipeline, we evaluated whether changes in each accelerometry measure were associated with disability outcomes or changes in brain atrophy measures (e.g., total brain and gray matter) by modeling within-person effects. To do so, we calculated an overall person-specific accelerometry mean (e.g., the average of given accelerometry measure for a person across all visits) and a within-person measure as the difference between the overall person-specific mean of a accelerometry measure and the person’s accelerometry measure at each time point. For disability outcomes (i.e., EDSS+, confirmed EDSS worsening, or 20% sustained worsening in T25FW), we used a Cox regression framework in which we included a term for the overall mean of a given measure as well as the time-varying within-person difference (Supplemental Methods) to evaluate their association with time to disability progression.^41–43^ Models were additionally adjusted for age, sex, race and BMI and were fit using a counting process style of input. For this analysis, only accelerometry measures collected prior to the detection of a confirmed disability worsening event were included in the calculation of within-person and overall person-specific means. For MRI models, we fit LME to account for within-person correlation and adjusted for a similar set of covariates as in disability models with the addition of EDSS. We also fit stratified models by HEAL-MS subgroup. Missing data for this study were minimal (<5% across all covariates) and were accounted for using indicator variables where applicable.

## Results

We included 239 participants who on average were 54.8 years (standard deviation; SD: 8.6 years), approximately 29% were male, and 23% were non-white races. With respect to MS characteristics, participants had an average MS duration of 13.2 years (SD: 8.2 years) with median baseline EDSS score 3.0 (IQR: 1.5); 43% were receiving an infusion therapy, and 34% had progressive MS. Full descriptive characteristics across the initial HEAL-MS groupings are provided in **Table 1**. Over an average of 2.9 (SD: 1.1) years for follow-up, 120 EDSS+ events occurred; initial analyses characterizing the cumulative incidence of disability progression also varied across HEAL-MS subgroups. People in the PMS subgroup tended to experience higher rates of disability accumulation (log-rank P for any difference in EDSS+ across groups=0.02; **Supplemental Figure 1A**). Results were also similar when we collapsed RRMS subgroups to use clinical groupings (log-rank for EDSS+ P=0.007; **Supplemental Figure 1B**).

**Table 1.** Demographic and clinical characteristics of included participants.

|  | Overall | HEAL-MS Subgroup |  |  |
| --- | --- | --- | --- | --- |
|  |  | RRMS<br>(stable) | RRMS<br>(suspected PMS) | PMS |
| N | 239 | 79 | 79 | 81 |
| N EDSS/MSFC assessments, mean (SD) | 6.03 (2.04) | 6.30 (1.69) | 5.92 (2.18) | 5.85 (2.21) |
| Disability outcome follow-up time, mean (SD), years | 2.89 (1.09) | 2.97 (0.74) | 2.94 (1.36) | 2.75 (1.07) |
| N MRIs, mean (SD) | 2.22 (1.02) | 2.20 (0.94) | 2.33 (1.09) | 2.14 (1.02) |
| MRI Follow-up time, mean (SD), years | 1.17 (0.93) | 1.13 (0.91) | 1.21 (0.96) | 1.16 (0.94) |
| Number of accelerometry wears, mean (SD) | 9.61 (3.97) | 9.59 (3.81) | 9.42 (4.27) | 9.80 (3.85) |
| Number of 24-hour periods (days) overall, mean (SD) | 86.07 (38.44) | 86.92 (37.20) | 84.20 (41.25) | 87.05 (37.18) |
| Number of days per wear, mean (SD) | 8.84 (1.07) | 8.91 (1.18) | 8.81 (1.09) | 8.80 (0.93) |
| log<sub>10</sub>(TAC) across all days, mean (SD) | 2096397.50<br>(646865.64) | 2206614.86<br>(511992.52) | 2195322.49<br>(699829.78) | 1892419.15<br>(669295.79) |
| Age (SD), years | 54.79 (8.61) | 52.94 (7.54) | 53.84 (8.40) | 57.53 (9.19) |
| Male sex at birth, n (%) | 69 (28.9) | 24 (30.4) | 19 (24.1) | 26 (32.1) |
| Race, n (%) |  |  |  |  |
| White | 184 (77.0) | 60 (75.9) | 60 (75.9) | 64 (79.0) |
| Black | 42 (17.6) | 17 (21.5) | 13 (16.5) | 12 (14.8) |
| Other races | 7 (2.9) | 0 (0.0) | 2 (2.5) | 5 (6.2) |
| Hispanic or Latinx ethnicity, n (%) | 9 (4.0) | 3 (3.9) | 3 (4.2) | 3 (3.8) |
| Years of education, mean (SD) | 13.99 (1.90) | 14.37 (1.32) | 13.75 (2.08) | 13.83 (2.16) |
| BMI, mean (SD), kg/m<sup>2</sup> | 28.60 (6.10) | 29.19 (6.11) | 28.40 (5.90) | 28.20 (6.30) |
| MS symptom duration, mean (SD), years | 13.24 (8.21) | 11.75 (7.22) | 13.79 (8.03) | 14.11 (9.12) |
| MS DMT, n (%) |  |  |  |  |
| Injectable interferon or glatiramer | 41 (17.2) | 12 (15.2) | 15 (19.0) | 14 (17.3) |
| Oral | 39 (16.3) | 19 (24.1) | 15 (19.0) | 5 (6.2) |
| Infusion or immune reconstitution | 103 (43.1) | 32 (40.5) | 39 (49.4) | 32 (39.5) |
| None | 56 (23.4) | 16 (20.3) | 10 (12.7) | 30 (37.0) |
| Baseline EDSS, median (IQR) | 3 (2.0, 4.5) | 2 (1.75-3.0) | 3 (2.0-3.5) | 5 (3.5-6.5) |

TAC: total activity count EDSS: Expanded Disability Status Scale MSFC: MS Functional Composite SD: standard deviation IQR: interquartile range

**Figure 1.**
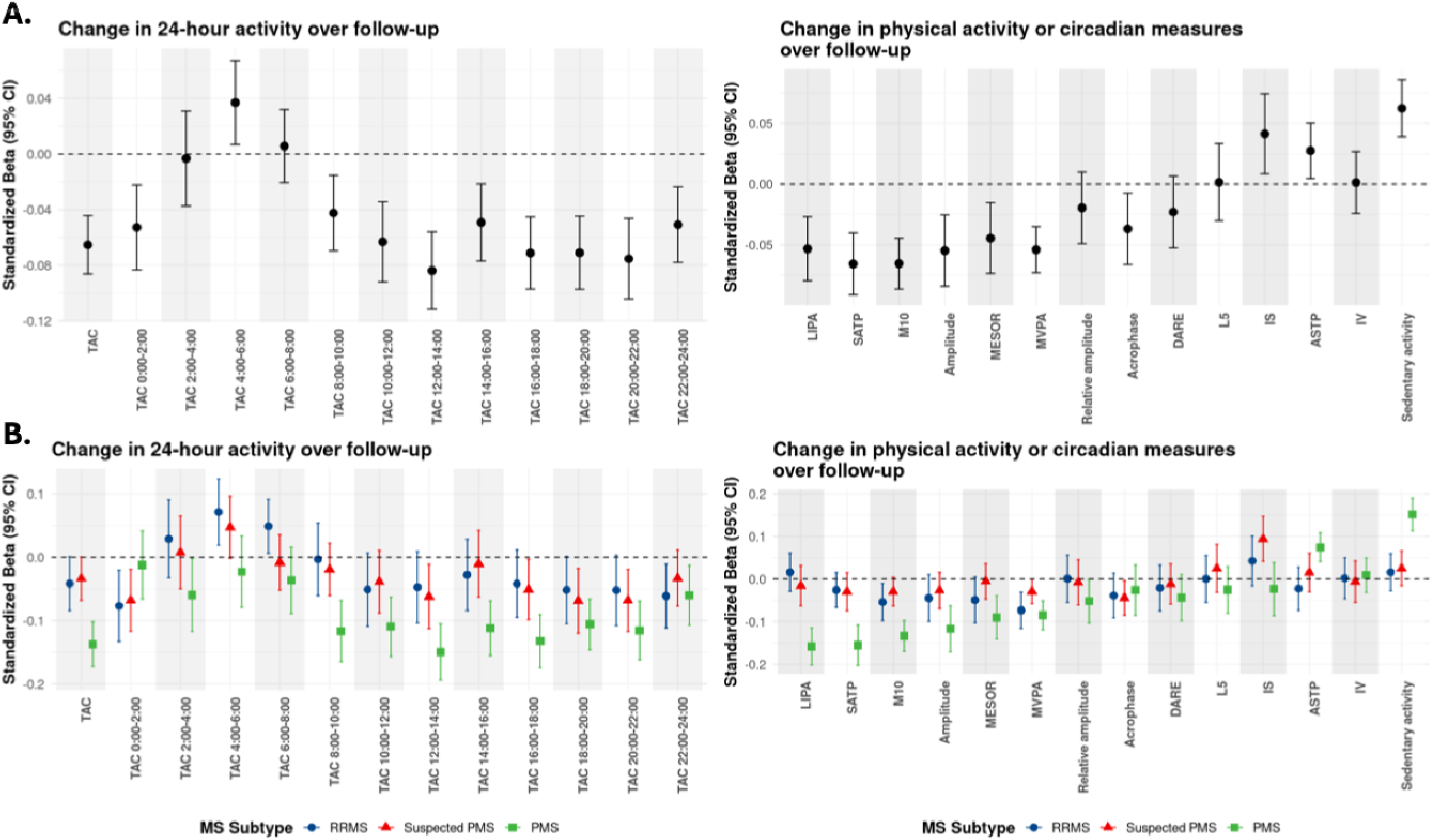
Change in accelerometry measures over follow-up overall and by subgroup. Figure 1. Annualized change in accelerometry values for 24-hour activity outcomes (left) and for physical activity and circadian parameters (right). Standardized beta coefficients and 95% CI are obtained from a linear mixed effects model adjusted for age, sex, race, and BMI. 1A includes all participants and 1B include estimates stratified by HEAL-MS subgroup (relapsing-remitting MS [RRMS], progressive MS (PMS), and suspected PMS.

### Changes in accelerometry measures over follow-up

Overall, participants wore accelerometers for an average of 9.6 (SD: 4.0) times, reflecting a total of 86.1 (SD: 38.4) days of wear over the approximate 3 years of follow-up. An average of 7.4 (SD: 3.6) wears for a mean 67.8 (SD: 34.4) total days of wear occurred prior to any disability progression events. We detected widespread changes in accelerometry measures that occurred over follow-up (**Supplemental Table 1**). **Figure 1A** shows standardized beta coefficients and 95% confidence intervals for changes in accelerometry-derived measures over follow-up (prior to any confirmed disability progression by EDSS+) and generally implicates a reduction in activity over time. Measures of overall activity (TAC and during 2-hour windows) and physical activity (MVPA and LIPA) generally decreased over follow-up whereas sedentary indicators (e.g., IS, sedentary activity) and measures of fragmentation (e.g., ASTP) tended to increase (**Figure 1** and **Supplemental Table 1**). For example, TAC decreased an average of 48,694 activity counts (roughly 0.10 SD of baseline levels) per year (95% CI: −33092, −64297; p=1.21×10^−9^). Circadian indices also changed during follow-up; relative amplitude decreased and M10 and MESOR tended to occur later in the day. Analyses were also consistent when adjusting for season (**Supplemental Figure 2**). When comparing how accelerometry indices changed between HEAL-MS subgroups, people with PMS exhibited more pronounced declines in multiple activity measures, including TAC, LIPA, and M10, compared to RRMS and suspected PMS (**Figure 1B** and **Supplemental Table 2**). In contrast, increases in sedentary activity and IV were most notable in the PMS group, with minimal changes observed in RRMS. These patterns suggest greater deterioration in activity and circadian rhythmicity among those with PMS. They were also generally similar when RRMS groups were collapsed into clinical subgroups (**Supplemental Figure 3**).

### Fluctuations in accelerometry measures and their translation to MS outcomes

#### Clinical outcomes

We chose to model the association for a 1 SD decrease in TAC/other accelerometry measures and risk of disability or MRI outcomes as, in general, accelerometry measures tended to decrease over time (e.g., **Figure 1**), and our goal was to test how these changes translate to clinical and imaging outcomes. We found that within-person decreases in daytime TAC (particularly 8:00-14:00) were consistently associated with higher risk of confirmed progression by EDSS+ (**Figure 2**), suggesting reductions in activity during specific time periods may precede progression detected by examination. For example, a 1 SD decrease in within-person (individual-level) activity from 8:00-10:00, 10:00-12:00 and 12:00-14:00 was associated with a respective 1.20 (95% CI: 1.04, 1.39; p=0.01), 1.22 (95% CI: 1.04, 1.41; p=0.008), and 1.23 (95% CI: 1.07, 1.42; p=0.007) higher risk of confirmed disability progression by EDSS+. These results were generally similar when considering individual EDSS+ components, including EDSS or 20% sustained worsening on T2FW. Notably, within-person decreases in TAC were not significantly associated with disability progression across outcomes (for EDSS+ HR: 1.14; 95% CI 0.96, 1.36; p=0.14). In exploratory analyses stratifying by HEAL-MS subgroup, results were generally similar, and we did not observe effect modification across the different groups (**Supplemental Figure 4**).

**Figure 2.**
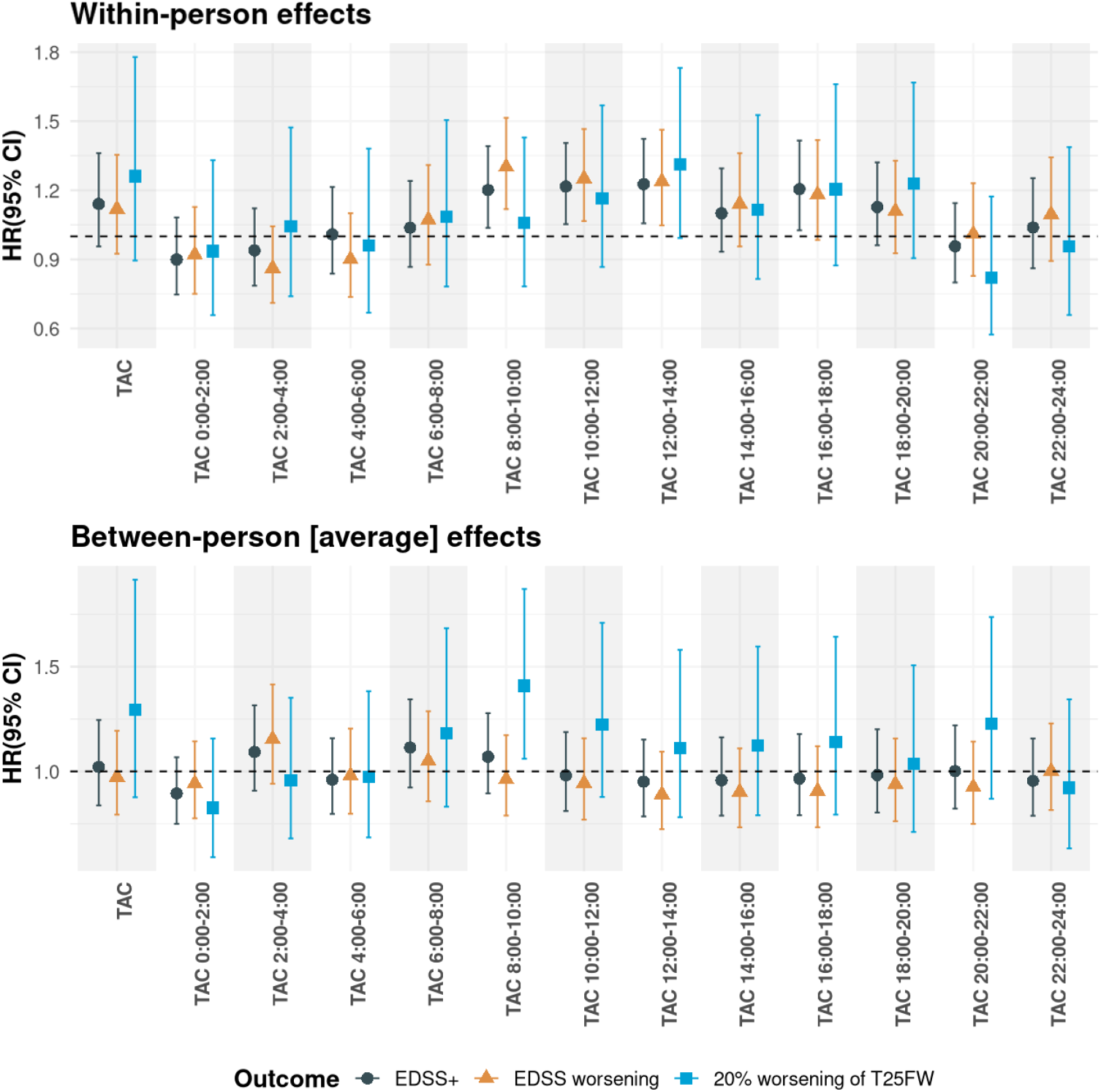
Effect estimates for a 1 SD reduction in overall and at specific time periods of the day and rates of subsequent disability progression for within and between (mean) measures. Figure 2. Within-person changes in total activity counts (TAC) or overall activity levels and risk of subsequent disability progression across different time intervals throughout the day. Hazard ratios (HRs) and 95% confidence intervals are shown for three disability outcomes: confirmed progression by EDSS+ (120 events), EDSS (98 events), and Timed 25-Foot Walk (T25FW; 33 events). Each HR reflects the association between a 1 SD decrease in activity overall or for a given period and risk of subsequent disability progression. A reference line at HR = 1 indicates no change in risk. The top panel reflects within-person changes (i.e., individual-level effects) as they correspond to risk whereas the bottom reflects the person-specific overall means (i.e., group-level effects) as they relate to risk. Effect estimates reflect a 1 SD decrease in TAC (and at specific time periods) as they relate to risk of EDSS+. We chose to model a 1 SD decrease as TAC generally decreased on average over follow-up.

For other activity measures, within-person reductions in ASTP (i.e., **Table 2**; less activity fragmentation) and MVPA (i.e., less physical activity) were associated with increased risk of disability progression (assessed by EDSS+). A 1 SD decrease in ASTP (i.e., less activity fragmentation over follow-up) was associated with a 12% lower risk of EDSS+-defined disability progression (HR: 0.88; 95% CI: 0.77–0.99; p = 0.04). A 1 SD decrease in MVPA (physical activity) was associated with a 28% higher risk (HR: 1.28; 95% CI: 1.04–1.56; p = 0.02). For circadian measures, a 1 SD decrease in DARE (daytime-to-overall activity ratio) was linked to a 25% increased risk of EDSS+ (HR: 1.25; 95% CI: 1.05–1.49; p = 0.01). A 1 SD decrease in IV was associated with a 16% lower risk (HR: 0.84; 95% CI: 0.72–0.98; p = 0.02). Between-person (mean) values for other activity and circadian measures showed similar patterns with changes in EDSS+. Findings were generally consistent in secondary analyses for EDSS and T25FW, though with wider confidence intervals due to fewer events.

**Table 2.** Hazard ratios for a 1 SD reduction* in other activity and circadian-based indices and disability progression.

|  | EDSS+<br>(120 events) |  |  |  | EDSS worsening<br>(98 events) |  |  |  | 20% T25FW worsening<br>(33 events) |  |  |  |
| --- | --- | --- | --- | --- | --- | --- | --- | --- | --- | --- | --- | --- |
| Effect type | Between |  | Within-person |  | Between |  | Within-person |  | Between |  | Within-person |  |
| HR 95% CI | HR (95% CI) | P | HR (95% CI) | P | HR (95% CI) | P | HR (95% CI) | P | HR (95% CI) | P | HR (95% CI) | P |
| **Physical activity measures** |  |  |  |  |  |  |  |  |  |  |  |  |
| ASTP | 1.00 (0.82, 1.22) | 0.99 | **0.88 (0.77, 0.99)** | **0.04** | 1.10 (0.88, 1.37) | 0.44 | **0.85 (0.74, 0.97)** | **0.02** | 0.78 (0.58, 1.06) | 0.11 | 0.93 (0.69, 1.25) | 0.64 |
| LIPA | 1.01 (0.84, 1.22) | 0.88 | 1.03 (0.87, 1.23) | 0.69 | 1.01 (0.84, 1.22) | 0.91 | 1.03 (0.85, 1.23) | 0.77 | 0.97 (0.69, 1.37) | 0.88 | 1.09 (0.78, 1.52) | 0.61 |
| MVPA | 1.06 (0.86, 1.30) | 0.57 | **1.28 (1.04, 1.56)** | **0.02** | 0.99 (0.8, 1.22) | 0.9 | 1.22 (0.98, 1.52) | 0.07 | **1.79 (1.08, 2.94)** | **0.02** | **1.61 (1.02, 2.5)** | **0.04** |
| SATP | 1.02 (0.84, 1.23) | 0.86 | 0.96 (0.8, 1.15) | 0.68 | 1.01 (0.83, 1.23) | 0.91 | 0.93 (0.75, 1.12) | 0.43 | 0.93 (0.66, 1.33) | 0.72 | 1.15 (0.82, 1.61) | 0.41 |
| Sedentary activity | 0.98 (0.81, 1.19) | 0.82 | 0.91 (0.76, 1.08) | 0.28 | 1.01 (0.83, 1.22) | 0.95 | 0.93 (0.77, 1.11) | 0.41 | 0.88 (0.61, 1.27) | 0.49 | 0.85 (0.61, 1.18) | 0.33 |
| **Circadian rhythm measures** |  |  |  |  |  |  |  |  |  |  |  |  |
| Acrophase | 0.91 (0.77, 1.06) | 0.24 | 0.99 (0.82, 1.19) | 0.93 | 0.95 (0.80, 1.12) | 0.55 | 0.97 (0.8, 1.19) | 0.81 | 0.79 (0.58, 1.08) | 0.14 | 1.06 (0.76, 1.49) | 0.72 |
| Amplitude | 0.89 (0.75, 1.05) | 0.18 | 0.98 (0.84, 1.15) | 0.84 | **0.85 (0.72, 0.99)** | **0.04** | 0.97 (0.83, 1.14) | 0.7 | 1.39 (0.93, 2.08) | 0.11 | 1.43 (0.98, 2.08) | 0.06 |
| DARE | 0.99 (0.82, 1.19) | 0.9 | **1.25 (1.05, 1.49)** | **0.01** | 0.92 (0.75, 1.12) | 0.39 | **1.32 (1.09, 1.59)** | **0.004** | 1.14 (0.83, 1.56) | 0.44 | 1.25 (0.89, 1.75) | 0.19 |
| IS | 1.00 (0.83, 1.20) | 0.97 | 0.85 (0.72, 1.01) | 0.07 | 0.98 (0.81, 1.19) | 0.86 | **0.82 (0.68, 0.99)** | **0.04** | 0.93 (0.65, 1.33) | 0.68 | 1.06 (0.75, 1.52) | 0.72 |
| IV | 1.00 (0.82, 1.22) | 0.97 | **0.84 (0.72, 0.98)** | **0.02** | 1.11 (0.90, 1.39) | 0.32 | **0.83 (0.70, 0.97)** | **0.02** | **0.64 (0.47, 0.88)** | **0.005** | 0.79 (0.62, 1.02) | 0.07 |
| L5 | 1.08 (0.88, 1.32) | 0.48 | 0.93 (0.77, 1.14) | 0.49 | 1.22 (0.97, 1.54) | 0.09 | 0.86 (0.70, 1.05) | 0.14 | 0.98 (0.69, 1.39) | 0.91 | 0.9 (0.65, 1.25) | 0.54 |
| M10 | 1.03 (0.84, 1.25) | 0.79 | 1.18 (0.99, 1.41) | 0.06 | 0.96 (0.78, 1.18) | 0.68 | 1.18 (0.97, 1.43) | 0.10 | 1.37 (0.92, 2.04) | 0.13 | 1.33 (0.96, 1.89) | 0.09 |
| MESOR | 0.93 (0.78, 1.11) | 0.42 | 0.96 (0.82, 1.12) | 0.63 | 0.90 (0.76, 1.06) | 0.22 | 0.93 (0.79, 1.10) | 0.39 | **1.52 (1.00, 2.33)** | **0.05** | **1.49 (1.03, 2.13)** | **0.03** |
| Relative amplitude | 0.92 (0.75, 1.12) | 0.42 | 1.18 (0.98, 1.41) | 0.08 | 0.79 (0.63, 1.0) | 0.05 | 1.3 (1.06, 1.59) | 0.009 | 1.14 (0.83, 1.56) | 0.42 | 1.25 (0.93, 1.69) | 0.14 |

Hazard ratios are represented as a 1 SD decrease in accelerometry measure to match how most measures changed over time (i.e., most measures decreased on average over follow-up). Thus, for within-person changes in MVPA, a 1 SD decrease in MVPA within an individual was associated with 28% increase in EDSS+. Likewise, a 1 SD decrease in fragmentation (which would be expected to be beneficial) is associated with a 12% reduction in risk of EDSS+. Values are all adjusted for age, sex, race and BMI
HR: hazard ratio
CI: confident interval
EDSS: Expanded Disability Status Scale
T25FW: timed 25-foot walk
ASTP: active to sedentary transition probability.
LIPA: low intensity physical activity
MVPA: moderate-to-vigorous physical activity
SATP: sedentary to active transition probability

### MRI

Within-person fluctuations in early morning activity (i.e., 6:00-10:00) were associated with differences in whole brain volume over time. For example, a 1 SD within-person decrease in TAC from 6:00-8:00 and 8:00-10:00 was associated with respective differences of −0.15% and −0.16% in whole brain volume over time (**Figure 3**; for 6:00-8:00: −0.15%; 95% CI: −0.003, −0.31; p=0.05; for 8:00-10:00:-0.16%; 95% CI: −0.04, −0.29; p=0.009). Decreasing activity within-person from 2:00-4:00 was also associated with higher whole brain volumes over time (per 1SD decrease: 0.14%; 95% CI: 0.02, 0.25; p=0.02). Findings were relatively consistent across 2-hour activity levels with respect to their relation to changes in deep gray matter and thalamic volumes for within-person models. A 1 SD decrease in activity from 8:00-10:00 was associated with −0.32% and −0.30% lower deep gray and thalamic volumes over time (deep gray: −0.32%; 95% CI: −0.13, −0.51; p=0.0009; thalamic: −0.30%; 95% CI: −0.52, −0.08; p=0.007). For mean models, higher total 24-hour activity as well as lower mean activity levels from 22:00-2:00 were associated positively with gray matter volumes over time (**Figure 2**).

**Figure 3.**
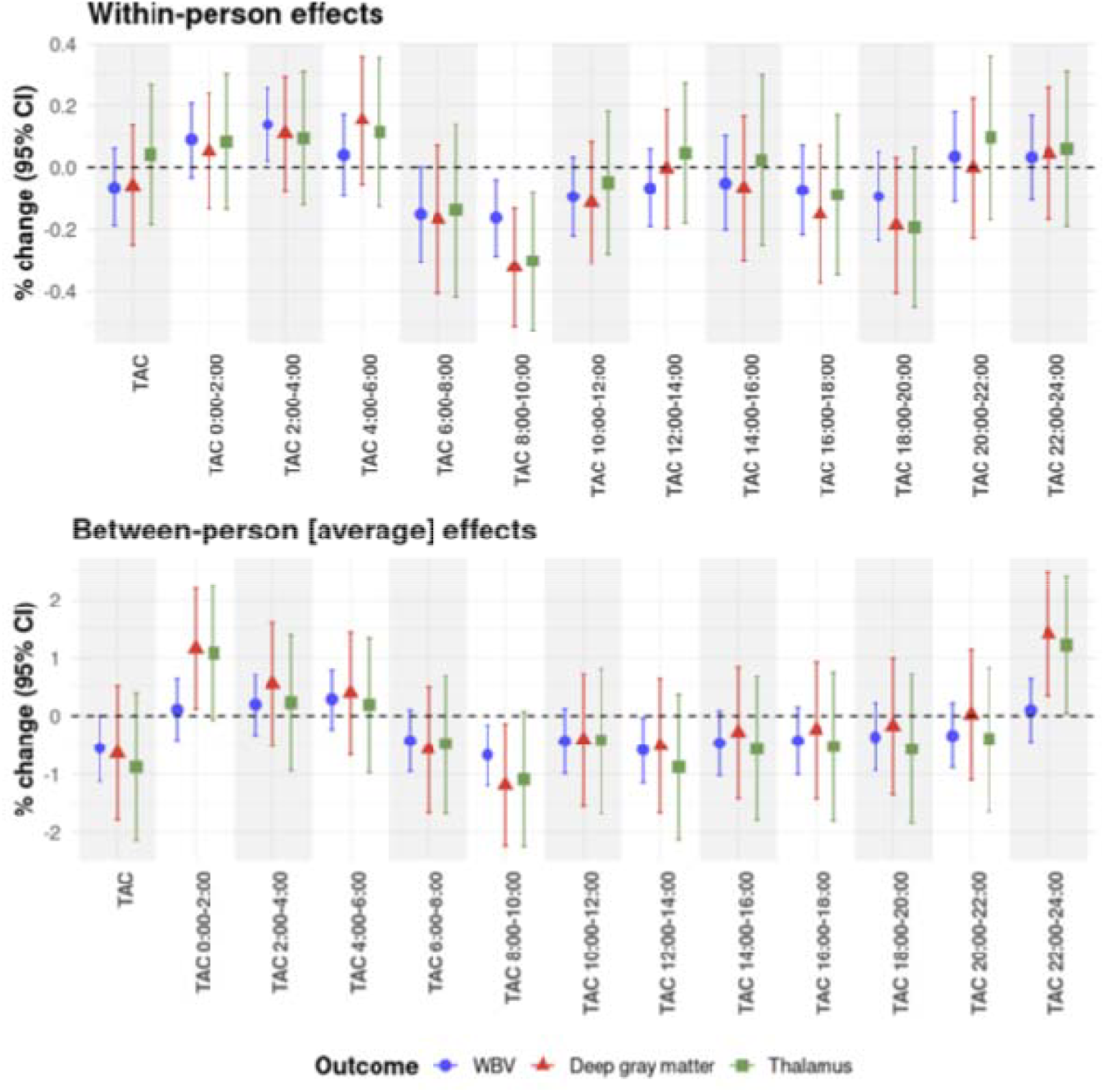
Effect estimates for a 1 SD reduction in overall and at specific time periods of the day and changes in MRI outcomes for within and between (mean) measures. Figure 3. Within-person changes in total activity counts (TAC) and mean TAC activity levels across different time intervals throughout the day and MRI outcomes. Effect estimates and 95% confidence intervals are shown for three MRI outcomes: whole brain volume (WBV), deep gray matter, and thalamic volume. Each estimates reflects the difference in brain volume for a 1 SD decrease in activity (either within person or in mean overall activity). A reference line at HR = 0 indicates no difference in brain volume. The top panel reflects within-person changes (i.e., individual-level effects) as they correspond to changes in brain volume whereas the bottom reflects the person-specific overall means (i.e., group-level effects) as they relate to brain volumes. Effect estimates reflect a 1 SD decrease in TAC (and at specific time periods) as they relate to brain volumes. We chose to model a 1 SD decrease as TAC generally decreased on average over follow-up.

For other activity and circadian measures, a 1 SD within-person decrease in DARE and relative amplitude (e.g., difference between active period and least active period) were associated with a respective loss of 0.26% and 0.18% in gray matter volume (**Table 3**; for DARE: −0.26%, 95%CI: −0.50, −0.03; p=0.03; for relative amplitude: 0.18%, 95% CI: −0.32, −0.04; p=0.01). For mean/between person models, lower average MVPA levels over follow-up were associated with worse brain and gray matter volumes over time (**Table 3**; per 1SD in average MVPA for whole brain volume: −0.81%; 95% CI: −0.24, −1.39; p=0.006; for deep matter volume: −1.34%; 95% CI: −0.17, −2.51; p=0.04; for thalamic volume: −1.59; 95% CI: −2.88, −0.31; p=0.02). Likewise, higher mean M10 (e.g., later timing of 10-hour activity) was associated with higher gray matter volumes over time (−0.63%; 95% CI: −0.05, −1.20; p=0.03). Other between-person and within-person accelerometry measures were not associated with MRI outcomes.

**Table 3.** Effect estimates for a 1 SD reduction* in other activity and circadian-based indices and brain volumes.

|  | WBV |  |  |  | Deep gray matter |  |  |  | Thalamus |  |  |  |
| --- | --- | --- | --- | --- | --- | --- | --- | --- | --- | --- | --- | --- |
| Effect type | Between |  | Within-person |  | Between |  | Within-person |  | Between |  | Within-person |  |
|  | Estimate<br>(95% CI) | P | Estimate<br>(95% CI) | P | Estimate<br>(95% CI) | P | Estimate<br>(95% CI) | P | Estimate<br>(95% CI) | P | Estimate<br>(95% CI) | P |
| **Physical activity measures** |  |  |  |  |  |  |  |  |  |  |  |  |
| ASTP | 0.27 (0.83, -0.29) | 0.34 | 0.04 (0.16, -0.08) | 0.49 | 0.04 (1.18, -1.10) | 0.94 | 0.09 (0.28, -0.09) | 0.33 | 0.04 (1.30, -1.22) | 0.95 | 0.10 (0.31, -0.12) | 0.39 |
| LIPA | -0.03 (0.49, -0.54) | 0.91 | -0.08 (0.04, -0.20) | 0.21 | 0.28 (1.33, -0.77) | 0.6 | -0.10 (0.09, -0.29) | 0.32 | 0.24 (1.40, -0.91) | 0.68 | -0.01 (0.21, -0.23) | 0.92 |
| MVPA | **-0.81 (-0.24, -1.39)** | **0.006** | -0.02 (0.12, -0.16) | 0.76 | **-1.34 (-0.17, -2.51)** | **0.03** | 0.03 (0.24, -0.18) | 0.78 | **-1.59 (-0.31, -2.88)** | **0.02** | 0.13 (0.38, -0.12) | 0.32 |
| SATP | -0.15 (0.37, -0.68) | 0.57 | -0.07 (0.05, -0.19) | 0.29 | 0.09 (1.16, -0.98) | 0.87 | 0.01 (0.19, -0.18) | 0.95 | -0.21 (0.97, -1.38) | 0.73 | 0.08 (0.30, -0.14) | 0.48 |
| Sedentary activity | 0.32 (0.86, -0.22) | 0.24 | 0.07 (0.19, -0.05) | 0.25 | 0.26 (1.37, -0.84) | 0.64 | 0.07 (0.26, -0.12) | 0.46 | 0.39 (1.60, -0.83) | 0.53 | -0.03 (0.20, -0.25) | 0.82 |
| **Circadian rhythm measures** |  |  |  |  |  |  |  |  |  |  |  |  |
| Acrophase | 0.28 (0.70, -0.14) | 0.19 | 0.08 (0.21, -0.06) | 0.26 | 0.79 (1.64, -0.06) | 0.07 | 0.05 (0.26, -0.15) | 0.61 | 0.82 (1.75, -0.12) | 0.09 | 0.02 (0.26, -0.22) | 0.86 |
| Amplitude | -0.28 (0.26, -0.81) | 0.31 | -0.03 (0.14, -0.20) | 0.73 | -0.65 (0.44, -1.74) | 0.24 | 0.08 (0.33, -0.18) | 0.55 | -0.74 (0.46, -1.93) | 0.23 | 0.18 (0.48, -0.12) | 0.24 |
| DARE | -0.22 (0.30, -0.75) | 0.41 | **-0.12 (-0.00, -0.25)** | **0.05** | -0.86 (0.21, -1.92) | 0.12 | **-0.24 (-0.05, -0.42)** | **0.02** | -0.62 (0.55, -1.80) | 0.30 | **-0.23 (-0.01, -0.45)** | **0.04** |
| IS | -0.06 (0.46, -0.58) | 0.82 | 0.07 (0.23, -0.08) | 0.34 | -0.64 (0.41, -1.69) | 0.24 | 0.12 (0.36, -0.12) | 0.32 | -0.45 (0.71, -1.61) | 0.45 | 0.19 (0.47, -0.09) | 0.18 |
| IV | 0.35 (0.92, -0.22) | 0.23 | 0.02 (0.14, -0.09) | 0.70 | 0.18 (1.34, -0.99) | 0.77 | 0.11 (0.29, -0.07) | 0.23 | 0.10 (1.38, -1.19) | 0.88 | 0.17 (0.38, -0.04) | 0.12 |
| L5 | 0.15 (0.65, -0.35) | 0.56 | **0.11 (0.22, -0.00)** | **0.05** | 0.11 (1.13, -0.91) | 0.83 | **0.22 (0.39, 0.06)** | **0.01** | 0.08 (1.20, -1.05) | 0.90 | 0.16 (0.35, -0.04) | 0.12 |
| M10 | **-0.63 (-0.05, -1.20)** | **0.03** | -0.06 (0.07, -0.20) | 0.35 | -0.87 (0.31, -2.05) | 0.15 | -0.08 (0.12, -0.29) | 0.44 | -1.10 (0.20, -2.40) | 0.10 | 0.02 (0.27, -0.22) | 0.86 |
| MESOR | -0.32 (0.20, -0.85) | 0.22 | 0.03 (0.20, -0.13) | 0.7 | -0.56 (0.50, -1.63) | 0.30 | 0.09 (0.35, -0.16) | 0.47 | -0.69 (0.48, -1.86) | 0.25 | 0.13 (0.43, -0.18) | 0.41 |
| Relative amplitude | -0.37 (0.17, -0.91) | 0.18 | **-0.18 (-0.04, -0.32)** | **0.01** | -0.37 (0.74, -1.48) | 0.51 | **-0.29 (-0.08, -0.51)** | **0.008** | -0.38 (0.84, -1.59) | 0.55 | -0.14 (0.12, -0.39) | 0.29 |

Effect estimates are represented as a 1 SD decrease in accelerometry measure to match how most measures changed over time (i.e., most measures decreased on average over follow-up). Thus, for between-person changes in MVPA, a 1 SD decrease in MVPA across individuals was associated with -0.81% lower brain volumes over time. Likewise, a 1 SD decrease in the daytime activity ratio (activity from 8:00-20:00 divided by the total activity from 0:00-23:59) which would reflect a dampening of daytime activity relative to activity over the course of a 24-hour period was associated with -0.12% lower brain volumes over time. Values are all adjusted for age, sex, race and BMI.
MRI: magnetic resonance imaging
WBV: whole brain volume
CI: confidence interval
ASTP: active to sedentary transition probability.
LIPA: low intensity physical activity
MVPA: moderate-to-vigorous physical activity
SATP: sedentary to active transition probability
DARE: activity from 8:00-20:00 divided by the total activity from 0:00-23:59

Findings were relatively consistent in analyses stratifying by HEAL-MS subgroup in that the observed associations between accelerometry measures (and within-person changes) were not modified in any of the categories (**Supplemental Table 4**).

## Discussion

In this relatively large longitudinal study of individuals with MS, we identified significant within-person changes in real-world physical activity and circadian rhythm patterns that preceded disability progression and brain atrophy. Declines in daytime activity, particularly from 8:00 to 14:00, and reductions in MVPA, DARE, and relative amplitude were consistently associated with greater risk of clinical worsening and loss of gray matter and thalamic volume. These associations were strongest among individuals with PMS, who showed more pronounced deterioration in accelerometry profiles than RRMS subgroups. Importantly, our ability to detect subclinical change prior to overt disability worsening highlights the value of longitudinal digital phenotyping for monitoring neurologic health and underscores the potential of accelerometry-derived metrics as prognostic tools. Notably, we detected associations over a relatively short time frame, supporting feasibility of detecting meaningful changes in activity patterns within periods relevant to Phase 2 trials for progressive MS.

Most accelerometry studies in MS have been cross-sectional. A recent systematic review identified 21 such studies comparing physical activity and sedentary time in PwMS versus controls.^18^ In general, cross-sectional studies show that (1) PwMS engage in less physical activity and have more sedentary time than age-matched healthy people, and (2) both physical activity and sedentary time correlate with disability levels, but in the opposite direction.^9–11,17,44^ For example, in a comparably-sized population of PwMS, Motl et al. correlated accelerometer output with walking behavior parameters and EDSS-defined disability levels in a cross-sectional analysis. We have shown that after adjusting for relevant covariates, lower activity counts are cross-sectionally correlated with progressive versus relapsing MS. For example, in a baseline analysis of the HEAL-MS cohort, we also found that lower overall activity as well as lower MVPA was correlated with disability levels Longitudinal studies in the field are sparse. Block et al., found that, in 95 PwMS, decreasing total step-count in a 1-year period was associated with worse clinic-based outcomes (timed 25-foot walk, timed-up-and-go, and patient-reported outcomes).^10^ Our finding that between-person differences in overall physical activity are associated with lower brain volumes corresponds nicely to the results from this study. We also build upon these results establishing widespread declines in activity measures derived using accelerometry as well as increases in markers of sedentary behavior. We further show how these changes in activity in specific time-periods translates to higher risk of subsequent disability worsening. That is, the effect estimates for changes in accelerometry measures over the course of the day that occur over follow-up (e.g., **Figure 1A**) mirror within-person associations for how decrements in activity over follow-up relate to higher risk of disability worsening and brain volume loss (i.e., top panels of **Figure 2** and **Figure 3**). These changes were detectable before confirmed disability worsening, supporting the predictive validity of accelerometry.

The relevant changes in accelerometry patterns may reflect occult neurodegeneration that later manifests as a change in brain volume and, ultimately, as disability worsening. However, it is noteworthy that in prior epidemiological studies of later-onset neurodegenerative disorders such as Alzheimer’s Disease, less physical activity and exercise in mid-life are associated with greater risk of subsequent disease incidence.^45^ Further, studies of aging have shown, using fairly standard accelerometry measures, that greater physical activity is associated longitudinally with brain volume preservation over several years.^46^ Whether the activity patterns herein that appear relevant to brain atrophy might also be a treatment target, such as by enhancing exercise or improving sleep quality, is certainly of interest. Our finding that increasing activity in in the night with a reduction in morning activities (suggestive of circadian disruption) is linked with more brain atrophy is also consistent with observations of circadian disruption in people with other neurodegenerative diseases (e.g., Alzheimer’s Disease, Parkinson’s Disease), in whom circadian disruption is commonly observed; research also suggests that these changes in patterns of activity manifest prior to the clinical manifestation or are potentially risk factors for each disease.^47^ Circadian disruption is also linked with cognitive decline in MCI.^48^

Strengths of this study include its novel design and inclusion of multiple accelerometry measures per person; each person contributed nearly 90 days of accelerometer wear-time (and 70 of which occurred prior to detection of clinical disability worsening). Other strengths include the inclusion of both overall activity and circadian patterns of activity, as many prior studies in MS have focused on total step count. We also considered a comprehensive set of circadian and physical activity measures and model changes at the individual rather than group level. This method is not only centered in precision medicine, which has implications for the development of more tailored treatment and monitoring approaches in MS, but within-person estimates also control for potential confounding variables that are static in an individual and but are not necessarily included in the model. Other strengths include the relatively large sample size (comparable to a Phase 2 trial in MS) compared with the existing longitudinal literature linking wearable devices in PwMS.

One limitation of this study includes the relatively short follow-up period for MRI, although if anything this would bias results toward the null, whereas we were able to detect significant loss of whole brain and gray matter volume loss during this period. Follow-up analyses will also include other imaging outcomes including those measuring spinal cord atrophy. Though hip-worn accelerometers, rather than wrist-worn for this cohort, may augment assessments of gait change, prior research in large-scale cohort studies (e.g., NHANES or UK Biobank) leveraged wrist-worn accelerometry. NHANES changed from hip-placed to wrist-worn devices to enhance participant adherence.^49^ Further, aspects of activity beyond gait may be relevant to predicting MS worsening and likewise will be more inclusive of individuals with MS who are no longer ambulatory. Finally, while activity patterns herein were characterized using research-grade accelerometers, which enhances the reliability of their detection, using these devices at scale (or clinic-wide) may not be feasible.

Future work should confirm findings in independent cohorts, define clinically meaningful thresholds for activity change, and examine links with cognition. Additional research is also needed leveraging commercial devices could improve scalability and allow assessment of other biomarkers like heart rate. Nonetheless, our findings demonstrate a compelling rationale for incorporating wearable accelerometers in MS clinical trials, particularly those focused on the prevention or reduction of MS disability.

## Data Availability

All data produced in the present study may be available upon reasonable request to the authors with the appropriate data sharing agreements in place.

## Funding

This study was supported by R01NR018851-01A1. Kathryn Fitzgerald is supported by K01MH121582-04 and TA-1805-31136 from the National MS Society. We also thank MS4MS for supporting our accelerometry program.

## Data availability

Data may be available to qualified investigators with the appropriate data sharing agreements in place.

## Competing Interests

Kathryn Fitzgerald receives consulting fees from SetPoint Medical. Ellen Mowry serves as PI for investigator-initiated studies from Biogen and Genentech. She also receives royalties for editorial duties on UpToDate. Peter Calabresi is PI on a grant from Genentech to JHU. He has received consulting fees from Lilly, Idorsia, and Novartis.

---

**Supplemental Figure 1.**
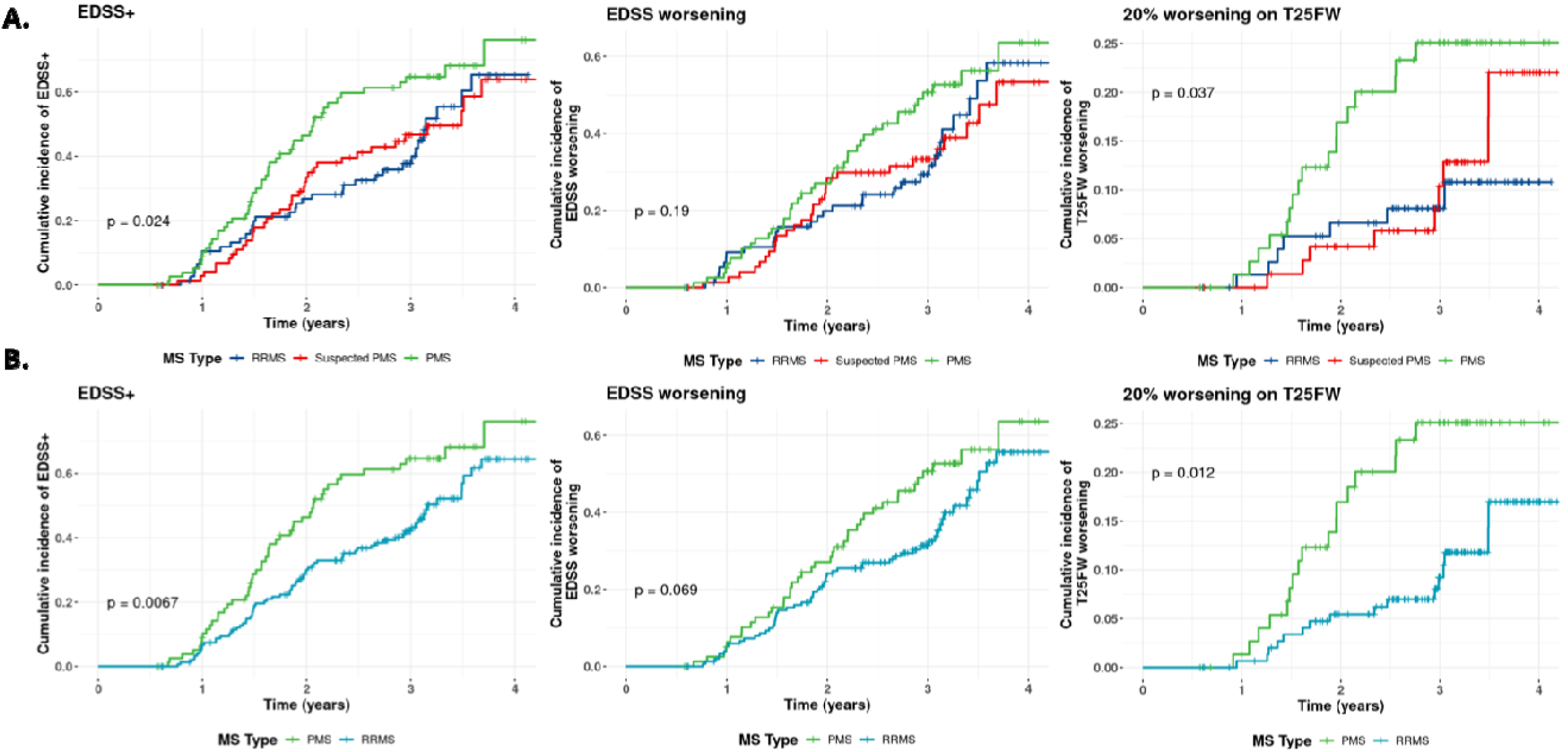
Cumulative incidence of disability progression across HEAL-MS subgroups and clinically defined subgroups. Supplemental Figure 1. Cumulative incidence curves are calculated as the complement of the Kaplan Meier (e.g., 1- [Kaplan Meier]) for each disability outcome: Expanded Disability Status Scale-plus (EDSS+; 120 events), confirmed EDSS worsening (98 events), and 20% worsening in Timed 25-Foot Walk (T25FW; 33 events) The p-value reflects the log-rank test. 1A. describes curves for HEAL-MS subgroups. 1B describes curves for clinically defined groups.

**Supplemental Figure 2.**
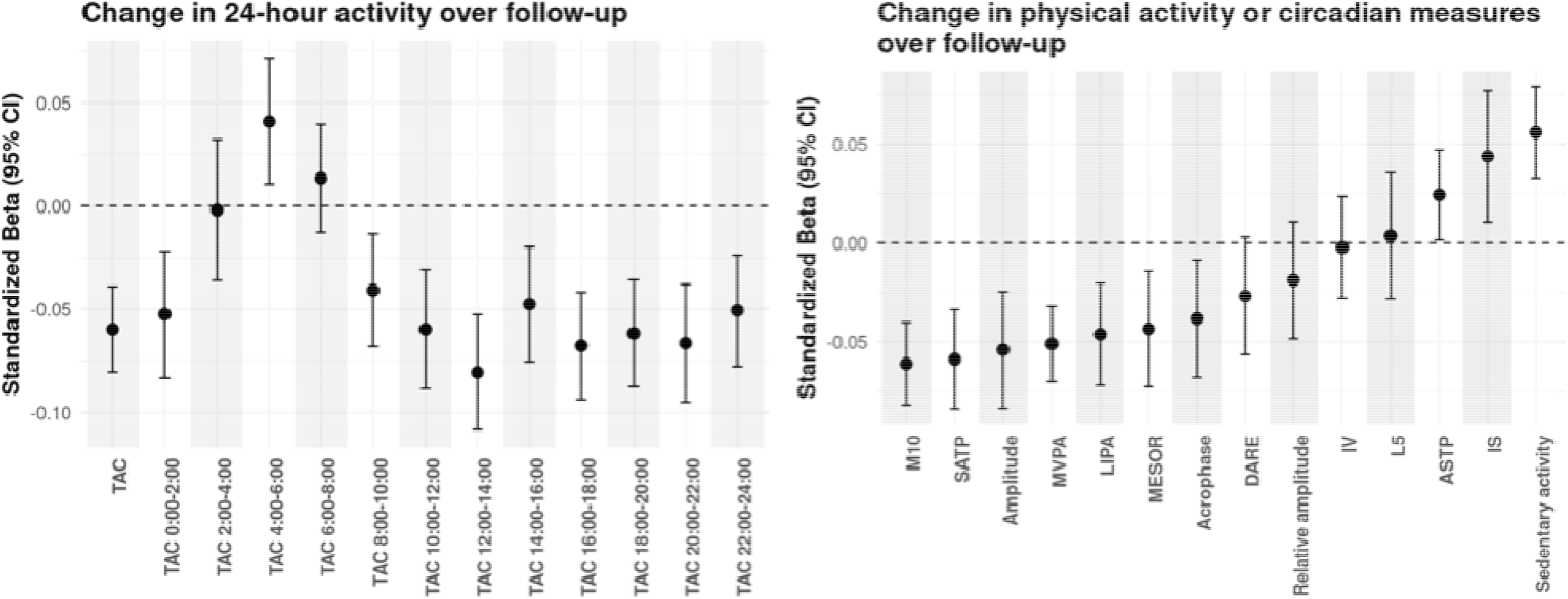
Changes in accelerometry measures over follow-up after adjusting for season. Supplemental Figure 2. Estimates are calculated from linear mixed effects models adjusted for age, sex, race and body mass index (BMI).

**Supplemental Figure 3.**
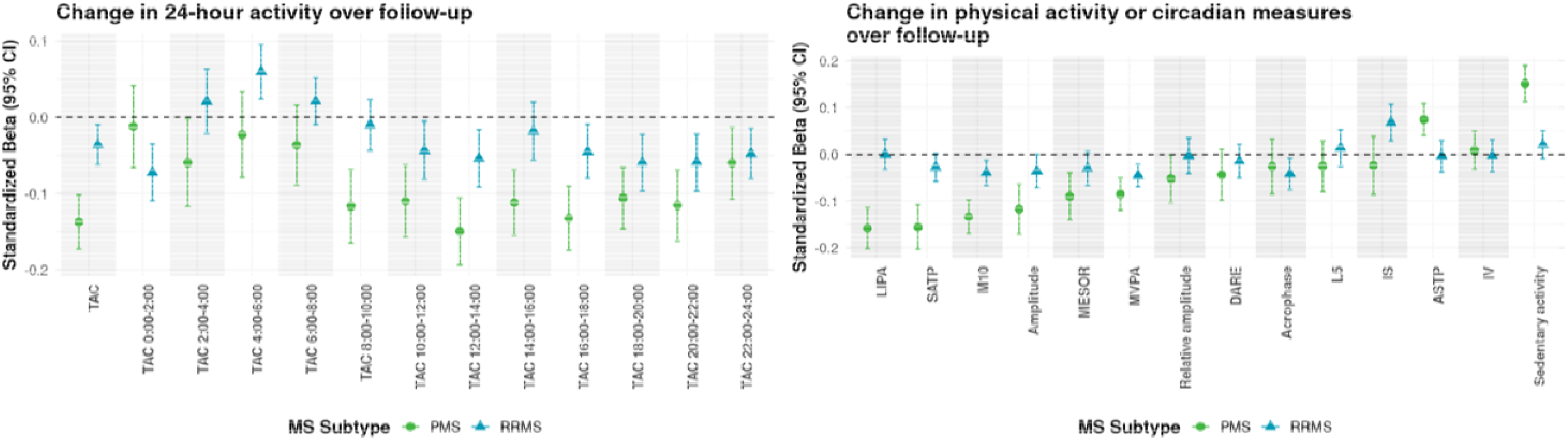
Change in accelerometry measures over follow-up by clinical subgroups (i.e., RRMS and PMS). Supplemental Figure 3. Estimates are calculated from linear mixed effects models adjusted for age, sex, race and body mass index (BMI).

**Supplemental Figure 4.**
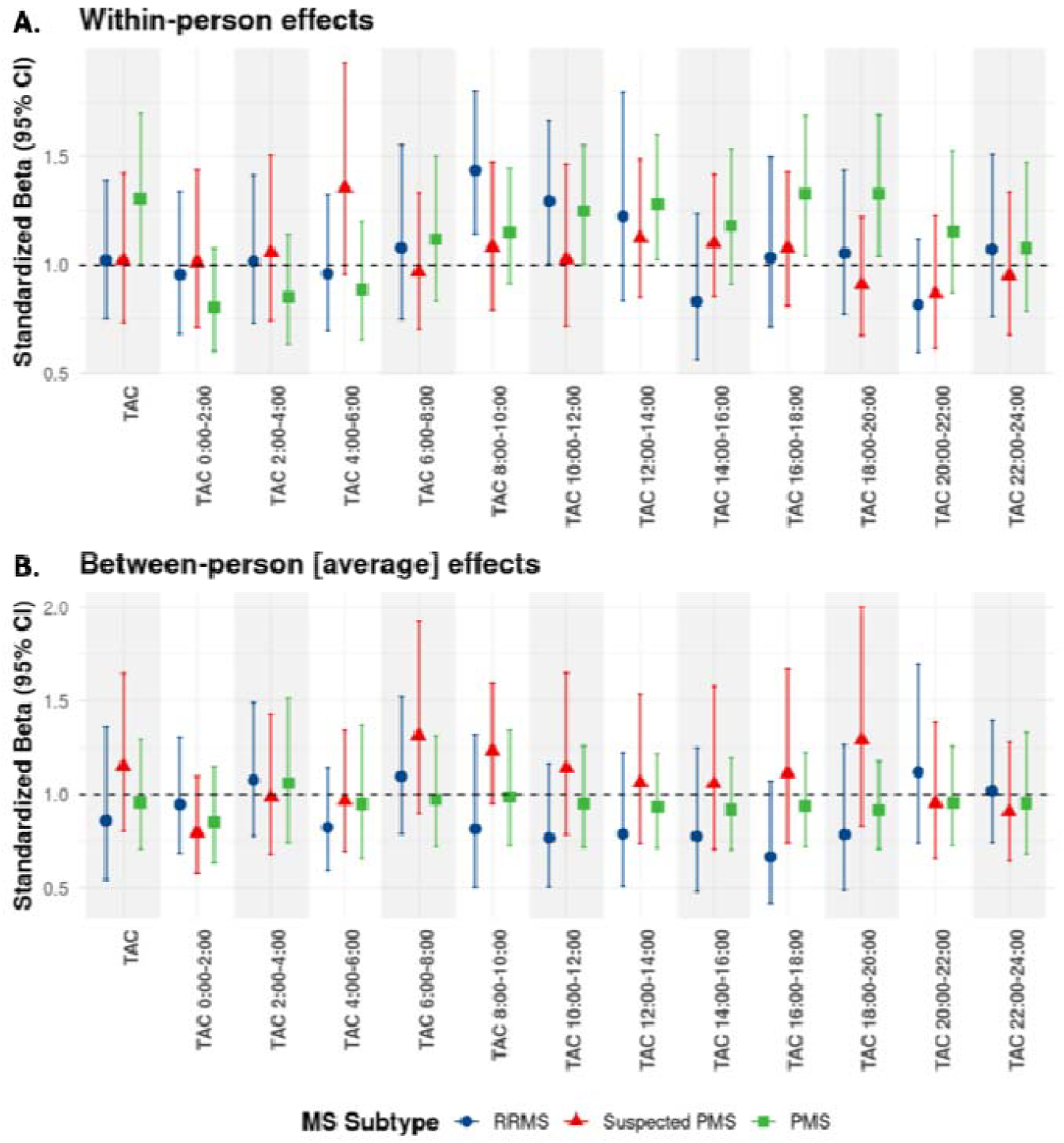
Effect estimates for a 1 SD reduction in overall and at specific time periods of the day and rates of EDSS+ for within and between (mean) measures by HEAL-MS-groups Supplemental Figure 4. Effect estimates reflect a 1 SD decrease in TAC (and at specific time periods) as they relate to risk of EDSS+. We chose to model a 1 SD decrease as TAC generally decreased on average over follow-up.

**Supplemental Table 1.**
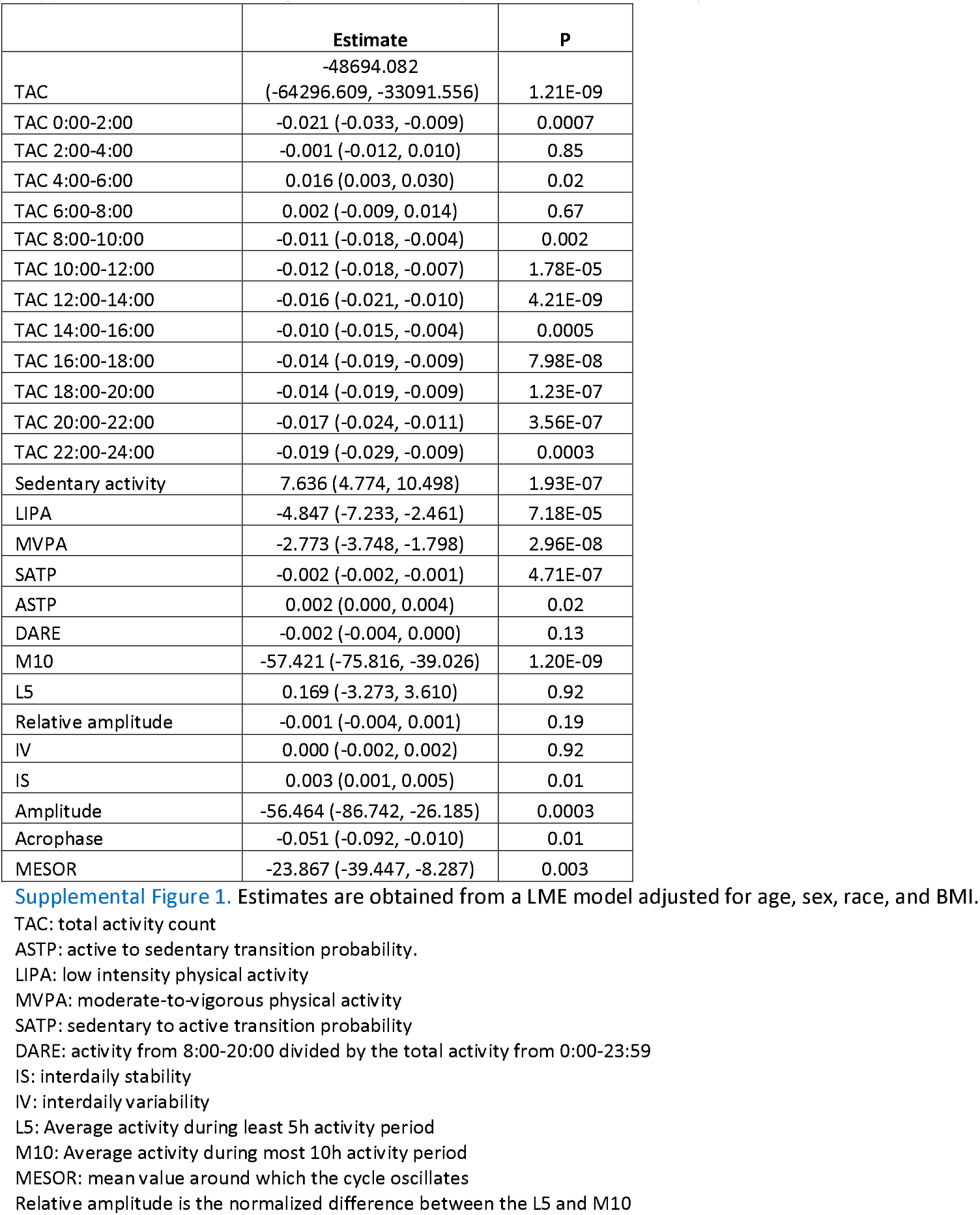
Change in accelerometry measures over follow-up.

**Supplemental Table 2.** Change in accelerometry measures over time stratified by HEAL-MS subgroup.

|  | HEAL-MS subgroup |  |  |  |  |  |  |  |
| --- | --- | --- | --- | --- | --- | --- | --- | --- |
|  | RRMS |  | Suspected PMS |  |  | PMS |  |  |
|  | Estimate<br>(95%CI) | P | Estimate<br>(95% CI) | P | P for RRMS<br>vs. sus PMS | Estimate<br>(95% CI) | P | P for RRMS<br>vs. PMS |
| TAC | -25044.607<br>(-50502.555, 413.341) | 0.05 | -25423.732<br>(-51779.875, 932.410) | 0.06 | 0.98 | -108710.409<br>(-137790.540, -79630.278) | 2.35E-13 | 2.34E-05 |
| TAC 0:00-2:00 | -0.028 (-0.048, -0.008) | 0.006 | -0.026 (-0.047, -0.006) | 0.01 | 0.91 | -0.005 (-0.028, 0.017) | 0.65 | 1.41E-01 |
| TAC 2:00-4:00 | 0.011 (-0.008, 0.029) | 0.26 | 0.003 (-0.016, 0.022) | 0.74 | 0.58 | -0.021 (-0.042, 0.000) | 0.05 | 2.85E-02 |
| TAC 4:00-6:00 | 0.032 (0.010, 0.054) | 0.004 | 0.022 (-0.001, 0.045) | 0.06 | 0.54 | -0.010 (-0.035, 0.015) | 0.44 | 1.42E-02 |
| TAC 6:00-8:00 | 0.021 (0.002, 0.040) | 0.03 | -0.003 (-0.022, 0.017) | 0.77 | 0.08 | -0.015 (-0.037, 0.006) | 0.16 | 1.30E-02 |
| TAC 8:00-10:00 | -0.001 (-0.012, 0.011) | 0.9 | -0.004 (-0.016, 0.008) | 0.48 | 0.67 | -0.033 (-0.046, -0.020) | 9.32E-07 | 3.25E-04 |
| TAC 10:00-12:00 | -0.009 (-0.018, 0.000) | 0.06 | -0.006 (-0.016, 0.003) | 0.21 | 0.69 | -0.025 (-0.036, -0.015) | 3.16E-06 | 2.32E-02 |
| TAC 12:00-14:00 | -0.007 (-0.016, 0.001) | 0.1 | -0.010 (-0.019, -0.001) | 0.02 | 0.63 | -0.034 (-0.043, -0.024) | 8.93E-12 | 5.77E-05 |
| TAC 14:00-16:00 | -0.004 (-0.013, 0.004) | 0.33 | -0.002 (-0.011, 0.007) | 0.71 | 0.68 | -0.026 (-0.037, -0.016) | 2.89E-07 | 1.31E-03 |
| TAC 16:00-18:00 | -0.006 (-0.014, 0.002) | 0.14 | -0.008 (-0.017, 0.000) | 0.05 | 0.72 | -0.030 (-0.040, -0.021) | 1.19E-10 | 1.17E-04 |
| TAC 18:00-20:00 | -0.008 (-0.017, 0.000) | 0.05 | -0.011 (-0.020, -0.002) | 0.01 | 0.66 | -0.025 (-0.034, -0.015) | 3.51E-07 | 1.11E-02 |
| TAC 20:00-22:00 | -0.011 (-0.022, -0.000) | 0.04 | -0.014 (-0.025, -0.003) | 0.01 | 0.72 | -0.029 (-0.042, -0.017) | 3.45E-06 | 3.14E-02 |
| TAC 22:00-24:00 | -0.022 (-0.038, -0.005) | 0.01 | -0.013 (-0.030, 0.004) | 0.13 | 0.48 | -0.022 (-0.040, -0.003) | 0.02 | 1.00E+00 |
| Sedentary activity | 1.705 (-2.945, 6.354) | 0.47 | 2.848 (-1.965, 7.662) | 0.25 | 0.74 | 21.342 (16.032, 26.653) | 3.33E-15 | 5.78E-08 |
| LIPA | 1.266 (-2.602, 5.134) | 0.52 | -1.234 (-5.237, 2.770) | 0.55 | 0.38 | -17.293 (-21.709, -12.878) | 1.64E-14 | 7.42E-10 |
| MVPA | -2.918 (-4.520, -1.317) | 0.0004 | -1.670 (-3.328, -0.012) | 0.05 | 0.29 | -3.979 (-5.809, -2.150) | 2.02E-05 | 3.93E-01 |
| SATP | -0.001 (-0.002, 0.000) | 0.25 | -0.001 (-0.002, 0.000) | 0.21 | 0.92 | -0.004 (-0.006, -0.003) | 4.47E-12 | 9.13E-06 |
| ASTP | -0.001 (-0.004, 0.002) | 0.42 | 0.001 (-0.002, 0.004) | 0.53 | 0.31 | 0.008 (0.005, 0.011) | 2.00E-06 | 4.15E-05 |
| DARE | -0.001 (-0.005, 0.002) | 0.4 | -0.001 (-0.004, 0.003) | 0.76 | 0.71 | -0.003 (-0.007, 0.001) | 0.1 | 4.88E-01 |
| M10 | -37.772 (-67.818, -7.725) | 0.01 | -26.144 (-57.249, 4.962) | 0.1 | 0.6 | -122.308 (-156.627, -87.990) | 2.84E-12 | 2.90E-04 |
| L5 | 0.198 (-5.461, 5.857) | 0.95 | 2.939 (-2.915, 8.793) | 0.33 | 0.51 | -2.999 (-9.449, 3.452) | 0.36 | 4.65E-01 |
| Relative amplitude | -0.000 (-0.004, 0.004) | 0.99 | -0.000 (-0.004, 0.003) | 0.81 | 0.87 | -0.005 (-0.009, -0.001) | 0.02 | 8.61E-02 |
| IV | 0.000 (-0.004, 0.004) | 0.92 | -0.000 (-0.004, 0.003) | 0.82 | 0.82 | 0.001 (-0.003, 0.005) | 0.68 | 8.05E-01 |
| IS | 0.003 (-0.001, 0.007) | 0.17 | 0.007 (0.003, 0.011) | 0.0007 | 0.14 | -0.002 (-0.006, 0.003) | 0.44 | 1.36E-01 |
| Amplitude | -50.692 (-100.421, -0.963) | 0.05 | -21.603 (-73.054, 29.849) | 0.41 | 0.43 | -109.108 (-165.822, -52.394) | 0.0002 | 1.29E-01 |
| Acrophase | -0.053 (-0.120, 0.015) | 0.13 | -0.059 (-0.129, 0.010) | 0.1 | 0.89 | -0.038 (-0.115, 0.039) | 0.34 | 7.76E-01 |
| MESOR | -30.137 (-55.726, -4.548) | 0.02 | -0.866 (-27.344, 25.613) | 0.95 | 0.12 | -44.852 (-74.044, -15.659) | 0.003 | 4.58E-01 |

**Supplemental Table 3.**
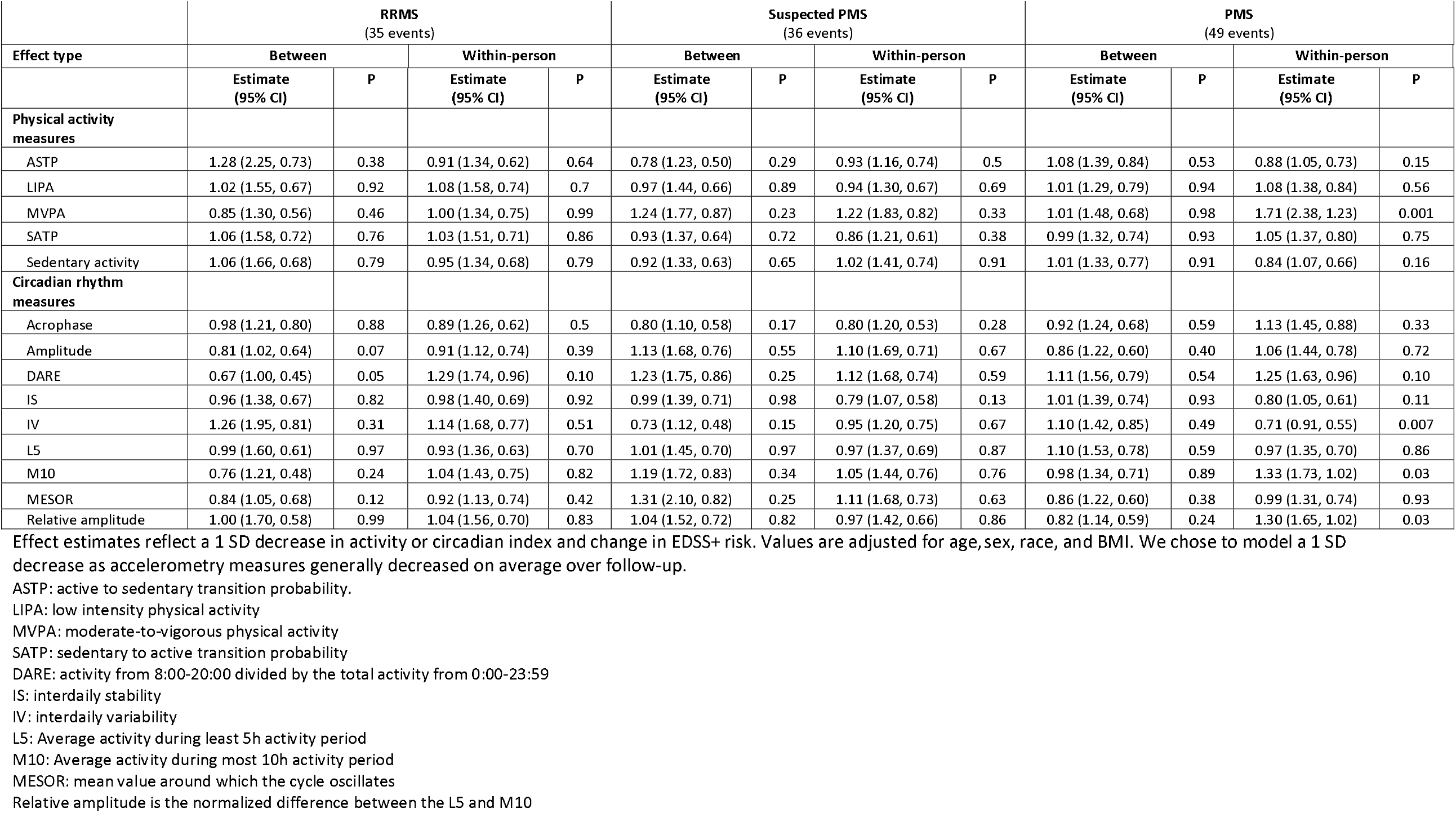
Effect estimates for a 1 SD reduction in other activity circadian-based indices and EDSS+ by HEAL-MS subgroups.

**Supplemental Table 4.**
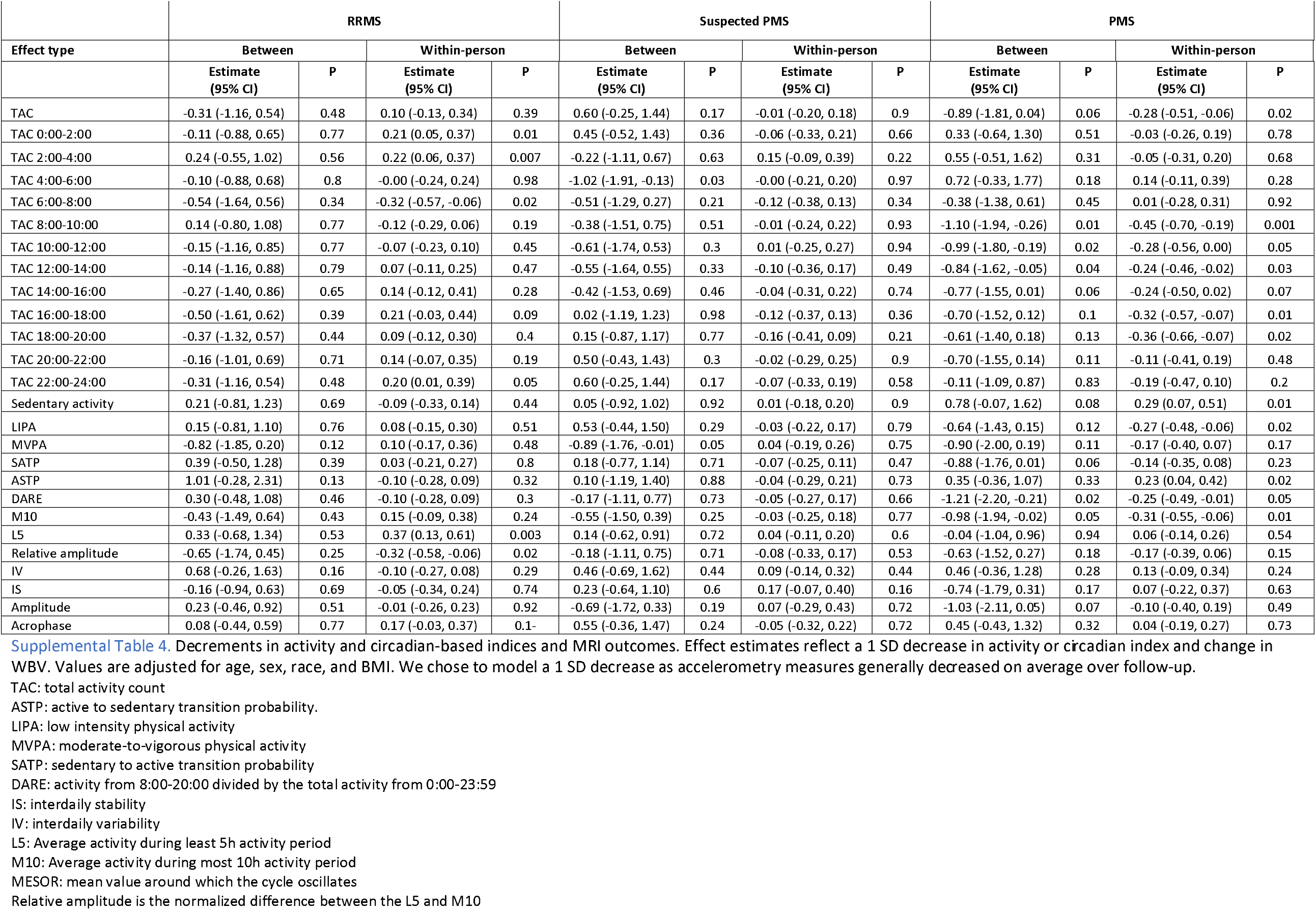
Effect estimates for a 1 SD reduction in activity and circadian-based indices and change in WBV by HEAL-MS subgroups.

|  | RRMS |  |  |  | Suspected PMS |  |  |  | PMS |  |  |  |
| --- | --- | --- | --- | --- | --- | --- | --- | --- | --- | --- | --- | --- |
| Effect type | Between |  | Within-person |  | Between |  | Within-person |  | Between |  | Within-person |  |
|  | Estimate<br>(95% CI) | P | Estimate<br>(95% CI) | P | Estimate<br>(95% CI) | P | Estimate<br>(95% CI) | P | Estimate<br>(95% CI) | P | Estimate<br>(95% CI) | P |
| TAC | -0.31 (-1.16, 0.54) | 0.48 | 0.10 (-0.13, 0.34) | 0.39 | 0.60 (-0.25, 1.44) | 0.17 | -0.01 (-0.20, 0.18) | 0.9 | -0.89 (-1.81, 0.04) | 0.06 | -0.28 (-0.51, -0.06) | 0.02 |
| TAC 0:00-2:00 | -0.11 (-0.88, 0.65) | 0.77 | 0.21 (0.05, 0.37) | 0.01 | 0.45 (-0.52, 1.43) | 0.36 | -0.06 (-0.33, 0.21) | 0.66 | 0.33 (-0.64, 1.30) | 0.51 | -0.03 (-0.26, 0.19) | 0.78 |
| TAC 2:00-4:00 | 0.24 (-0.55, 1.02) | 0.56 | 0.22 (0.06, 0.37) | 0.007 | -0.22 (-1.11, 0.67) | 0.63 | 0.15 (-0.09, 0.39) | 0.22 | 0.55 (-0.51, 1.62) | 0.31 | -0.05 (-0.31, 0.20) | 0.68 |
| TAC 4:00-6:00 | -0.10 (-0.88, 0.68) | 0.8 | -0.00 (-0.24, 0.24) | 0.98 | -1.02 (-1.91, -0.13) | 0.03 | -0.00 (-0.21, 0.20) | 0.97 | 0.72 (-0.33, 1.77) | 0.18 | 0.14 (-0.11, 0.39) | 0.28 |
| TAC 6:00-8:00 | -0.54 (-1.64, 0.56) | 0.34 | -0.32 (-0.57, -0.06) | 0.02 | -0.51 (-1.29, 0.27) | 0.21 | -0.12 (-0.38, 0.13) | 0.34 | -0.38 (-1.38, 0.61) | 0.45 | 0.01 (-0.28, 0.31) | 0.92 |
| TAC 8:00-10:00 | 0.14 (-0.80, 1.08) | 0.77 | -0.12 (-0.29, 0.06) | 0.19 | -0.38 (-1.51, 0.75) | 0.51 | -0.01 (-0.24, 0.22) | 0.93 | -1.10 (-1.94, -0.26) | 0.01 | -0.45 (-0.70, -0.19) | 0.001 |
| TAC 10:00-12:00 | -0.15 (-1.16, 0.85) | 0.77 | -0.07 (-0.23, 0.10) | 0.45 | -0.61 (-1.74, 0.53) | 0.3 | 0.01 (-0.25, 0.27) | 0.94 | -0.99 (-1.80, -0.19) | 0.02 | -0.28 (-0.56, 0.00) | 0.05 |
| TAC 12:00-14:00 | -0.14 (-1.16, 0.88) | 0.79 | 0.07 (-0.11, 0.25) | 0.47 | -0.55 (-1.64, 0.55) | 0.33 | -0.10 (-0.36, 0.17) | 0.49 | -0.84 (-1.62, -0.05) | 0.04 | -0.24 (-0.46, -0.02) | 0.03 |
| TAC 14:00-16:00 | -0.27 (-1.40, 0.86) | 0.65 | 0.14 (-0.12, 0.41) | 0.28 | -0.42 (-1.53, 0.69) | 0.46 | -0.04 (-0.31, 0.22) | 0.74 | -0.77 (-1.55, 0.01) | 0.06 | -0.24 (-0.50, 0.02) | 0.07 |
| TAC 16:00-18:00 | -0.50 (-1.61, 0.62) | 0.39 | 0.21 (-0.03, 0.44) | 0.09 | 0.02 (-1.19, 1.23) | 0.98 | -0.12 (-0.37, 0.13) | 0.36 | -0.70 (-1.52, 0.12) | 0.1 | -0.32 (-0.57, -0.07) | 0.01 |
| TAC 18:00-20:00 | -0.37 (-1.32, 0.57) | 0.44 | 0.09 (-0.12, 0.30) | 0.4 | 0.15 (-0.87, 1.17) | 0.77 | -0.16 (-0.41, 0.09) | 0.21 | -0.61 (-1.40, 0.18) | 0.13 | -0.36 (-0.66, -0.07) | 0.02 |
| TAC 20:00-22:00 | -0.16 (-1.01, 0.69) | 0.71 | 0.14 (-0.07, 0.35) | 0.19 | 0.50 (-0.43, 1.43) | 0.3 | -0.02 (-0.29, 0.25) | 0.9 | -0.70 (-1.55, 0.14) | 0.11 | -0.11 (-0.41, 0.19) | 0.48 |
| TAC 22:00-24:00 | -0.31 (-1.16, 0.54) | 0.48 | 0.20 (0.01, 0.39) | 0.05 | 0.60 (-0.25, 1.44) | 0.17 | -0.07 (-0.33, 0.19) | 0.58 | -0.11 (-1.09, 0.87) | 0.83 | -0.19 (-0.47, 0.10) | 0.2 |
| Sedentary activity | 0.21 (-0.81, 1.23) | 0.69 | -0.09 (-0.33, 0.14) | 0.44 | 0.05 (-0.92, 1.02) | 0.92 | 0.01 (-0.18, 0.20) | 0.9 | 0.78 (-0.07, 1.62) | 0.08 | 0.29 (0.07, 0.51) | 0.01 |
| LIPA | 0.15 (-0.81, 1.10) | 0.76 | 0.08 (-0.15, 0.30) | 0.51 | 0.53 (-0.44, 1.50) | 0.29 | -0.03 (-0.22, 0.17) | 0.79 | -0.64 (-1.43, 0.15) | 0.12 | -0.27 (-0.48, -0.06) | 0.02 |
| MVPA | -0.82 (-1.85, 0.20) | 0.12 | 0.10 (-0.17, 0.36) | 0.48 | -0.89 (-1.76, -0.01) | 0.05 | 0.04 (-0.19, 0.26) | 0.75 | -0.90 (-2.00, 0.19) | 0.11 | -0.17 (-0.40, 0.07) | 0.17 |
| SATP | 0.39 (-0.50, 1.28) | 0.39 | 0.03 (-0.21, 0.27) | 0.8 | 0.18 (-0.77, 1.14) | 0.71 | -0.07 (-0.25, 0.11) | 0.47 | -0.88 (-1.76, 0.01) | 0.06 | -0.14 (-0.35, 0.08) | 0.23 |
| ASTP | 1.01 (-0.28, 2.31) | 0.13 | -0.10 (-0.28, 0.09) | 0.32 | 0.10 (-1.19, 1.40) | 0.88 | -0.04 (-0.29, 0.21) | 0.73 | 0.35 (-0.36, 1.07) | 0.33 | 0.23 (0.04, 0.42) | 0.02 |
| DARE | 0.30 (-0.48, 1.08) | 0.46 | -0.10 (-0.28, 0.09) | 0.3 | -0.17 (-1.11, 0.77) | 0.73 | -0.05 (-0.27, 0.17) | 0.66 | -1.21 (-2.20, -0.21) | 0.02 | -0.25 (-0.49, -0.01) | 0.05 |
| M10 | -0.43 (-1.49, 0.64) | 0.43 | 0.15 (-0.09, 0.38) | 0.24 | -0.55 (-1.50, 0.39) | 0.25 | -0.03 (-0.25, 0.18) | 0.77 | -0.98 (-1.94, -0.02) | 0.05 | -0.31 (-0.55, -0.06) | 0.01 |
| L5 | 0.33 (-0.68, 1.34) | 0.53 | 0.37 (0.13, 0.61) | 0.003 | 0.14 (-0.62, 0.91) | 0.72 | 0.04 (-0.11, 0.20) | 0.6 | -0.04 (-1.04, 0.96) | 0.94 | 0.06 (-0.14, 0.26) | 0.54 |
| Relative amplitude | -0.65 (-1.74, 0.45) | 0.25 | -0.32 (-0.58, -0.06) | 0.02 | -0.18 (-1.11, 0.75) | 0.71 | -0.08 (-0.33, 0.17) | 0.53 | -0.63 (-1.52, 0.27) | 0.18 | -0.17 (-0.39, 0.06) | 0.15 |
| IV | 0.68 (-0.26, 1.63) | 0.16 | -0.10 (-0.27, 0.08) | 0.29 | 0.46 (-0.69, 1.62) | 0.44 | 0.09 (-0.14, 0.32) | 0.44 | 0.46 (-0.36, 1.28) | 0.28 | 0.13 (-0.09, 0.34) | 0.24 |
| IS | -0.16 (-0.94, 0.63) | 0.69 | -0.05 (-0.34, 0.24) | 0.74 | 0.23 (-0.64, 1.10) | 0.6 | 0.17 (-0.07, 0.40) | 0.16 | -0.74 (-1.79, 0.31) | 0.17 | 0.07 (-0.22, 0.37) | 0.63 |
| Amplitude | 0.23 (-0.46, 0.92) | 0.51 | -0.01 (-0.26, 0.23) | 0.92 | -0.69 (-1.72, 0.33) | 0.19 | 0.07 (-0.29, 0.43) | 0.72 | -1.03 (-2.11, 0.05) | 0.07 | -0.10 (-0.40, 0.19) | 0.49 |
| Acrophase | 0.08 (-0.44, 0.59) | 0.77 | 0.17 (-0.03, 0.37) | 0.1- | 0.55 (-0.36, 1.47) | 0.24 | -0.05 (-0.32, 0.22) | 0.72 | 0.45 (-0.43, 1.32) | 0.32 | 0.04 (-0.19, 0.27) | 0.73 |

TAC: total activity count
ASTP: active to sedentary transition probability.
LIPA: low intensity physical activity
MVPA: moderate-to-vigorous physical activity
SATP: sedentary to active transition probability
DARE: activity from 8:00-20:00 divided by the total activity from 0:00-23:59
IS: interdaily stability
IV: interdaily variability
L5: Average activity during least 5h activity period

## Notes

### Author Declarations

Ethics committee/IRB of Johns Hopkins University gave ethical approval for this work

### Summary of Updates

We included updated follow-up to include more eligible participants as well as now have included clinical outcomes.

