## Supplemental Methods for "Short-term changes in objectively measured activity predict brain atrophy and disability progression in multiple sclerosis"

Measures derived from accelerometry are dynamic over follow-up and the translation of how within-person changes correspond to disability or atrophy outcomes is a critical component of the advancement of precision medicine in MS. specifically, modeling of within-person or individual-level effects can inform early intervention or advance personalized disease monitoring strategies. This analytic structure is also advantageous as it allows us to account for unmeasured fixed factors of an individual that would otherwise confound estimates across individuals. Individual-level effects were of particular interest, so we designed our analysis to be able to evaluate both within-person (individual) and between-person (group or average) level effects. This strategy was employed through the following steps (data is not actual data but is for demonstration purposes only):

1. Creation of a between-person effect. The MEAN_TAC is obtained as the average TAC for all accelerometry visits (prior to EDSS+) for all individuals

| **ID** | **TAC at Time 1** | **TAC at Time 2** | **TAC at Time 3** | **TAC at Time 4** | **MEAN TAC** |
| --- | --- | --- | --- | --- | --- |
| 1 | 1049471 | 1418031 | 1371714 | 1474240 | 1328364 |
| 2 | 1135820 | 1392765 | 1284711 | 1423953 | 1309312 |
| 3 | 1063422 | 1451932 | 1368207 | 1510285 | 1348462 |
| 4 | 1027593 | 1382176 | 1329408 | 1448610 | 1296947 |
| 5 | 1092317 | 1426033 | 1351010 | 1467219 | 1334145 |

2. The within-person variable is then calculated as the difference between a person’s activity at that specific time point from that person’s mean activity (e.g., for ID 1 for Time 1: 1049471-1328364=-278893).

| **ID** | **TAC for Time 1**  **within** | **TAC for Time 2**  **within** | **TAC for Time 3**  **within** | **TAC for Time 4**  **within** |
| --- | --- | --- | --- | --- |
| 1 | -278893 | 89667 | 43350 | 145876 |
| 2 | -173492 | 83453 | -24601 | 114641 |
| 3 | -285040 | 103470 | 19745 | 161823 |
| 4 | -269354 | 85229 | 32461 | 151663 |
| 5 | -241828 | 91888 | 16865 | 133074 |

3. The wide dataset is then transformed into a long-format dataset for longitudinal analysis.

| **ID** | **Time** | **TAC_mean** | **TAC_within** |
| --- | --- | --- | --- |
| 1 | 1 | 1328364 | -278893 |
| 1 | 2 | 1328364 | 89667 |
| 1 | 3 | 1328364 | 43350 |
| 1 | 4 | 1328364 | 145876 |
| 2 | 1 | 1309312 | -173492 |
| 2 | 2 | 1309312 | 83453 |
| 2 | 3 | 1309312 | -24601 |
| 2 | 4 | 1309312 | 114641 |
| 3 | 1 | 1348462 | -285040 |
| 3 | 2 | 1348462 | 103470 |
| 3 | 3 | 1348462 | 19745 |
| 3 | 4 | 1348462 | 161823 |
| 4 | 1 | 1296947 | -269354 |
| 4 | 2 | 1296947 | 85229 |
| 4 | 3 | 1296947 | 32461 |
| 4 | 4 | 1296947 | 151663 |
| 5 | 1 | 1334145 | -241828 |
| 5 | 2 | 1334145 | 91888 |
| 5 | 3 | 1334145 | 16865 |
| 5 | 4 | 1334145 | 133074 |

The regression model then includes both the TAC_mean and TAC_within. The coefficients are interpreted as a one-unit change in TAC for the within or between person effects. For the purposes of our manuscript, each was standardized to reflect a 1 SD unit difference.
